# Japan COVID-19 Task Force: a nation-wide consortium to elucidate host genetics of COVID-19 pandemic in Japan

**DOI:** 10.1101/2021.05.17.21256513

**Authors:** Ho Namkoong, Ryuya Edahiro, Koichi Fukunaga, Yuya Shirai, Kyuto Sonehara, Hiromu Tanaka, Ho Lee, Takanori Hasegawa, Masahiro Kanai, Tatsuhiko Naito, Kenichi Yamamoto, Ryunosuke Saiki, Takayoshi Hyugaji, Eigo Shimizu, Kotoe Katayama, Kazuhisa Takahashi, Norihiro Harada, Toshio Naito, Makoto Hiki, Yasushi Matsushita, Haruhi Takagi, Ryousuke Aoki, Ai Nakamura, Sonoko Harada, Hitoshi Sasano, Hiroki Kabata, Katsunori Masaki, Hirofumi Kamata, Shinnosuke Ikemura, Shotaro Chubachi, Satoshi Okamori, Hideki Terai, Atsuho Morita, Takanori Asakura, Junichi Sasaki, Hiroshi Morisaki, Yoshifumi Uwamino, Kosaku Nanki, Yohei Mikami, Sho Uchida, Shunsuke Uno, Rino Ishihara, Yuta Matsubara, Tomoyasu Nishimura, Takanori Ogawa, Takashi Ishiguro, Taisuke Isono, Shun Shibata, Yuma Matsui, Chiaki Hosoda, Kenji Takano, Takashi Nishida, Yoichi Kobayashi, Yotaro Takaku, Noboru Takayanagi, Soichiro Ueda, Ai Tada, Masayoshi Miyawaki, Masaomi Yamamoto, Eriko Yoshida, Reina Hayashi, Tomoki Nagasaka, Sawako Arai, Yutaro Kaneko, Kana Sasaki, Etsuko Tagaya, Masatoshi Kawana, Ken Arimura, Kunihiko Takahashi, Tatsuhiko Anzai, Satoshi Ito, Akifumi Endo, Yuji Uchimura, Yasunari Miyazaki, Takayuki Honda, Tomoya Tateishi, Shuji Tohda, Naoya Ichimura, Kazunari Sonobe, Chihiro Sassa, Jun Nakajima, Yasushi Nakano, Yukiko Nakajima, Ryusuke Anan, Ryosuke Arai, Yuko Kurihara, Yuko Harada, Kazumi Nishio, Tetsuya Ueda, Masanori Azuma, Ryuichi Saito, Toshikatsu Sado, Yoshimune Miyazaki, Ryuichi Sato, Yuki Haruta, Tadao Nagasaki, Yoshinori Yasui, Yoshinori Hasegawa, Yoshikazu Mutoh, Tomonori Sato, Reoto Takei, Satoshi Hagimoto, Yoichiro Noguchi, Yasuhiko Yamano, Hajime Sasano, Sho Ota, Yasushi Nakamori, Kazuhisa Yoshiya, Fukuki Saito, Tomoyuki Yoshihara, Daiki Wada, Hiromu Iwamura, Syuji Kanayama, Shuhei Maruyama, Takashi Yoshiyama, Ken Ohta, Hiroyuki Kokuto, Hideo Ogata, Yoshiaki Tanaka, Kenichi Arakawa, Masafumi Shimoda, Takeshi Osawa, Hiroki Tateno, Isano Hase, Shuichi Yoshida, Shoji Suzuki, Miki Kawada, Hirohisa Horinouchi, Fumitake Saito, Keiko Mitamura, Masao Hagihara, Junichi Ochi, Tomoyuki Uchida, Rie Baba, Daisuke Arai, Takayuki Ogura, Hidenori Takahashi, Shigehiro Hagiwara, Genta Nagao, Shunichiro Konishi, Ichiro Nakachi, Koji Murakami, Mitsuhiro Yamada, Hisatoshi Sugiura, Hirohito Sano, Shuichiro Matsumoto, Nozomu Kimura, Yoshinao Ono, Hiroaki Baba, Yusuke Suzuki, Sohei Nakayama, Keita Masuzawa, Shinichi Namba, Ken Suzuki, Nobuyuki Hizawa, Takayuki Shiroyama, Satoru Miyawaki, Yusuke Kawamura, Akiyoshi Nakayama, Hirotaka Matsuo, Yuichi Maeda, Takuro Nii, Yoshimi Noda, Takayuki Niitsu, Yuichi Adachi, Takatoshi Enomoto, Saori Amiya, Reina Hara, Toshihiro Kishikawa, Shuhei Yamada, Shuhei Kawabata, Noriyuki Kijima, Masatoshi Takagaki, Noa Sasa, Yuya Ueno, Motoyuki Suzuki, Norihiko Takemoto, Hirotaka Eguchi, Takahito Fukusumi, Takao Imai, Munehisa Fukushima, Haruhiko Kishima, Hidenori Inohara, Kazunori Tomono, Kazuto Kato, Meiko Takahashi, Fumihiko Matsuda, Haruhiko Hirata, Yoshito Takeda, Hidefumi Koh, Tadashi Manabe, Yohei Funatsu, Fumimaro Ito, Takahiro Fukui, Keisuke Shinozuka, Sumiko Kohashi, Masatoshi Miyazaki, Tomohisa Shoko, Mitsuaki Kojima, Tomohiro Adachi, Motonao Ishikawa, Kenichiro Takahashi, Takashi Inoue, Toshiyuki Hirano, Keigo Kobayashi, Hatsuyo Takaoka, Kazuyoshi Watanabe, Naoki Miyazawa, Yasuhiro Kimura, Reiko Sado, Hideyasu Sugimoto, Akane Kamiya, Naota Kuwahara, Akiko Fujiwara, Tomohiro Matsunaga, Yoko Sato, Takenori Okada, Yoshihiro Hirai, Hidetoshi Kawashima, Atsuya Narita, Kazuki Niwa, Yoshiyuki Sekikawa, Koichi Nishi, Masaru Nishitsuji, Mayuko Tani, Junya Suzuki, Hiroki Nakatsumi, Takashi Ogura, Hideya Kitamura, Eri Hagiwara, Kota Murohashi, Hiroko Okabayashi, Takao Mochimaru, Shigenari Nukaga, Ryosuke Satomi, Yoshitaka Oyamada, Nobuaki Mori, Tomoya Baba, Yasutaka Fukui, Mitsuru Odate, Shuko Mashimo, Yasushi Makino, Kazuma Yagi, Mizuha Hashiguchi, Junko Kagyo, Tetsuya Shiomi, Satoshi Fuke, Hiroshi Saito, Tomoya Tsuchida, Shigeki Fujitani, Mumon Takita, Daiki Morikawa, Toru Yoshida, Takehiro Izumo, Minoru Inomata, Naoyuki Kuse, Nobuyasu Awano, Mari Tone, Akihiro Ito, Yoshihiko Nakamura, Kota Hoshino, Junichi Maruyama, Hiroyasu Ishikura, Tohru Takata, Toshio Odani, Masaru Amishima, Takeshi Hattori, Yasuo Shichinohe, Takashi Kagaya, Toshiyuki Kita, Kazuhide Ohta, Satoru Sakagami, Kiyoshi Koshida, Kentaro Hayashi, Tetsuo Shimizu, Yutaka Kozu, Hisato Hiranuma, Yasuhiro Gon, Namiki Izumi, Kaoru Nagata, Ken Ueda, Reiko Taki, Satoko Hanada, Kodai Kawamura, Kazuya Ichikado, Kenta Nishiyama, Hiroyuki Muranaka, Kazunori Nakamura, Naozumi Hashimoto, Keiko Wakahara, Sakamoto Koji, Norihito Omote, Akira Ando, Nobuhiro Kodama, Yasunari Kaneyama, Shunsuke Maeda, Takashige Kuraki, Takemasa Matsumoto, Koutaro Yokote, Taka-Aki Nakada, Ryuzo Abe, Taku Oshima, Tadanaga Shimada, Masahiro Harada, Takeshi Takahashi, Hiroshi Ono, Toshihiro Sakurai, Takayuki Shibusawa, Yoshifumi Kimizuka, Akihiko Kawana, Tomoya Sano, Chie Watanabe, Ryohei Suematsu, Hisako Sageshima, Ayumi Yoshifuji, Kazuto Ito, Saeko Takahashi, Kota Ishioka, Morio Nakamura, Makoto Masuda, Aya Wakabayashi, Hiroki Watanabe, Suguru Ueda, Masanori Nishikawa, Yusuke Chihara, Mayumi Takeuchi, Keisuke Onoi, Jun Shinozuka, Atsushi Sueyoshi, Yoji Nagasaki, Masaki Okamoto, Sayoko Ishihara, Masatoshi Shimo, Yoshihisa Tokunaga, Yu Kusaka, Takehiko Ohba, Susumu Isogai, Aki Ogawa, Takuya Inoue, Satoru Fukuyama, Yoshihiro Eriguchi, Akiko Yonekawa, Keiko Kan-o, Koichiro Matsumoto, Kensuke Kanaoka, Shoichi Ihara, Kiyoshi Komuta, Yoshiaki Inoue, Shigeru Chiba, Kunihiro Yamagata, Yuji Hiramatsu, Hirayasu Kai, Koichiro Asano, Tsuyoshi Oguma, Yoko Ito, Satoru Hashimoto, Masaki Yamasaki, Yu Kasamatsu, Yuko Komase, Naoya Hida, Takahiro Tsuburai, Baku Oyama, Minoru Takada, Hidenori Kanda, Yuichiro Kitagawa, Tetsuya Fukuta, Takahito Miyake, Shozo Yoshida, Shinji Ogura, Shinji Abe, Yuta Kono, Yuki Togashi, Hiroyuki Takoi, Ryota Kikuchi, Shinichi Ogawa, Tomouki Ogata, Shoichiro Ishihara, Arihiko Kanehiro, Shinji Ozaki, Yasuko Fuchimo, Sae Wada, Nobukazu Fujimoto, Kei Nishiyama, Mariko Terashima, Satoru Beppu, Kosuke Yoshida, Osamu Narumoto, Hideaki Nagai, Nobuharu Ooshima, Mitsuru Motegi, Akira Umeda, Kazuya Miyagawa, Hisato Shimada, Mayu Endo, Yoshiyuki Ohira, Masafumi Watanabe, Sumito Inoue, Akira Igarashi, Masamichi Sato, Hironori Sagara, Akihiko Tanaka, Shin Ohta, Tomoyuki Kimura, Yoko Shibata, Yoshinori Tanino, Takefumi Nikaido, Hiroyuki Minemura, Yuki Sato, Yuichiro Yamada, Takuya Hashino, Masato Shinoki, Hajime Iwagoe, Hiroshi Takahashi, Kazuhiko Fujii, Hiroto Kishi, Masayuki Kanai, Tomonori Imamura, Tatsuya Yamashita, Masakiyo Yatomi, Toshitaka Maeno, Shinichi Hayashi, Mai Takahashi, Mizuki Kuramochi, Isamu Kamimaki, Yoshiteru Tominaga, Tomoo Ishii, Mitsuyoshi Utsugi, Akihiro Ono, Toru Tanaka, Takeru Kashiwada, Kazue Fujita, Yoshinobu Saito, Masahiro Seike, Yosuke Omae, Yasuhito Nannya, Takafumi Ueno, Tomomi Takano, Kazuhiko Katayama, Masumi Ai, Atsushi Kumanogoh, Toshiro Sato, Naoki Hasegawa, Katsushi Tokunaga, Makoto Ishii, Ryuji Koike, Yuko Kitagawa, Akinori Kimura, Seiya Imoto, Satoru Miyano, Seishi Ogawa, Takanori Kanai, Yukinori Okada

**Affiliations:** Department of Infectious Diseases, Keio University School of Medicine, Tokyo, Japan; Department of Statistical Genetics, Osaka University Graduate School of Medicine, Suita, Japan; Department of Respiratory Medicine and Clinical Immunology, Osaka University Graduate School of Medicine, Suita, Japan; Division of Pulmonary Medicine, Department of Medicine, Keio University School of Medicine, Tokyo, Japan; Integrated Frontier Research for Medical Science Division, Institute for Open and Transdisciplinary Research Initiatives, Osaka University, Suita, Japan; M&D Data Science Center, Tokyo Medical and Dental University, Tokyo, Japan; Department of Biomedical Informatics, Harvard Medical School, Boston, MA, USA; Department of Pathology and Tumor Biology, Kyoto University, Kyoto, Japan; Division of Health Medical Intelligence, Human Genome Center, the Institute of Medical Science, the University of Tokyo, Tokyo, Japan; Department of Respiratory Medicine, Juntendo University Faculty of Medicine and Graduate School of Medicine, Tokyo, Japan; Department of General Medicine, Juntendo University Faculty of Medicine and Graduate School of Medicine, Tokyo, Japan; Department of Emergency and Disaster Medicine, Juntendo University Faculty of Medicine and Graduate School of Medicine, Tokyo, Japan; Department of Cardiovascular Biology and Medicine, Juntendo University Faculty of Medicine and Graduate School of Medicine, Tokyo, Japan; Department of Internal Medicine and Rheumatology, Juntendo University Faculty of Medicine and Graduate School of Medicine, Tokyo, Japan; Department of Nephrology, Juntendo University Faculty of Medicine and Graduate School of Medicine, Tokyo, Japan; Atopy (Allergy) Research Center, Juntendo University Graduate School of Medicine, Tokyo, Japan; Department of Emergency and Critical Care Medicine, Keio University School of Medicine, Tokyo, Japan; Department of Anesthesiology, Keio University School of Medicine, Tokyo, Japan; Department of Laboratory Medicine, Keio University School of Medicine, Tokyo, Japan; Division of Gastroenterology and Hepatology, Department of Medicine, Keio University School of Medicine, Tokyo, Japan; Keio University Health Center, Tokyo, Japan; Department of Respiratory Medicine, Saitama Cardiovascular and Respiratory Center, Kumagaya, Japan; JCHO (Japan Community Health care Organization) Saitama Medical Center, Internal Medicine, Saitama, Japan; Department of Respiratory Medicine, Tokyo Women’s Medical University, Tokyo, Japan; Department of General Medicine, Tokyo Women’s Medical University, Tokyo, Japan; Clinical Research Center, Tokyo Medical and Dental University Hospital of Medicine, Tokyo, Japan; Department of Medical Informatics, Tokyo Medical and Dental University Hospital of Medicine, Tokyo, Japan; Respiratory Medicine, Tokyo Medical and Dental University, Tokyo, Japan; Clinical Laboratory, Tokyo Medical and Dental University Hospital of Medicine, Tokyo, Japan; Kawasaki Municipal Ida Hospital, Department of Internal Medicine, Kawasaki, Japan; Department of Respiratory Medicine, Osaka Saiseikai Nakatsu Hospital, Osaka, Japan; Department of Infection Control, Osaka Saiseikai Nakatsu Hospital, Osaka, Japan; Department of Infectious Diseases, Tosei General Hospital, Seto, Japan; Department of Respiratory, Allergic Diseases Internal Medicine, Tosei General Hospital, Seto, Japan; Department of Emergency and Critical Care Medicine, Kansai Medical University General Medical Center, Moriguchi, Japan; Fukujuji hospital, Kiyose, Japan; Department of Pulmonary Medicine, Saitama City Hospital, Saitama, Japan; Department of Infectious Diseases, Saitama City Hospital, Saitama, Japan; Department of General Thoracic Surgery, Saitama City Hospital, Saitama, Japan; Department of Pulmonary Medicine, Eiju General Hospital, Tokyo, Japan; Division of Infection Control, Eiju General Hospital, Tokyo, Japan; Department of Hematology, Eiju General Hospital, Tokyo, Japan; Saiseikai Utsunomiya Hospital, Utsunomiya, Japan; Department of Respiratory Medicine, Tohoku University Graduate School of Medicine, Sendai, Japan; Department of Infectious Diseases, Tohoku University Graduate School of Medicine, Sendai, Japan; Department of Respiratory Medicine, Kitasato University Kitasato Institute Hospital, Tokyo, Japan; Department of Pulmonary Medicine, Faculty of Medicine, University of Tsukuba, Tsukuba, Japan; Department of Neurosurgery, Faculty of Medicine, the University of Tokyo, Tokyo, Japan; Department of Integrative Physiology and Bio-Nano Medicine, National Defense Medical College, Tokorozawa, Japan; Department of Otorhinolaryngology-Head and Neck Surgery, Osaka University Graduate School of Medicine, Suita, Japan; Department of Head and Neck Surgery, Aichi Cancer Center Hospital, Nagoya, Japan; Department of Neurosurgery, Osaka University Graduate School of Medicine, Suita, Japan; Department of Otolaryngology and Head and Neck Surgery, Kansai Rosai Hospital, Hyogo, Japan; Division of Infection Control and Prevention, Osaka University Hospital, Suita, Japan; Department of Biomedical Ethics and Public Policy, Osaka University Graduate School of Medicine, Suita, Japan; Center for Genomic Medicine, Kyoto University Graduate School of Medicine, Kyoto, Japan; Tachikawa Hospital, Tachikawa, Japan; Department of Emergency and Critical Care Medicine, Tokyo Women’s Medical University Medical Center East, Tokyo, Japan; Department of Medicine, Tokyo Women’s Medical University Medical Center East, Tokyo, Japan; Department of Pediatrics, Tokyo Women’s Medical University Medical Center East, Tokyo, Japan; Internal Medicine, Sano Kosei General Hospital, Sano, Japan; Japan Community Health care Organization Kanazawa Hospital, Kanazawa, Japan; Department of Respiratory Medicine, Saiseikai Yokohamashi Nanbu Hospital, Yokohama, Japan; Department of Clinical Laboratory, Saiseikai Yokohamashi Nanbu Hospital, Yokohama, Japan; Internal Medicine, Internal Medicine Center, Showa University Koto Toyosu Hospital, Tokyo, Japan; Department of Respiratory Medicine, Japan Organization of Occupational Health and Safety, Kanto Rosai Hospital, Kawasaki, Japan; Department of General Internal Medicine, Japan Organization of Occupational Health and Safety, Kanto Rosai Hospital, Kawasaki, Japan; Ishikawa Prefectural Central Hospital, Kanazawa, Japan; Kanagawa Cardiovascular and Respiratory Center, Yokohama, Japan; Department of Respiratory Medicine, National Hospital Organization Tokyo Medical Center, Tokyo, Japan; Department of Allergy, National Hospital Organization Tokyo Medical Center, Tokyo, Japan; Department of General Internal Medicine and Infectious Diseases, National Hospital Organization Tokyo Medical Center, Tokyo, Japan; Department of Respiratory Medicine, Toyohashi Municipal Hospital, Toyohashi, Japan; Keiyu Hospital, Yokohama, Japan; KKR Sapporo Medical Center, Department of respiratory medicine, Sapporo, Japan; Division of General Internal Medicine, Department of Internal Medicine, St. Marianna University School of Medicine, Kawasaki, Japan; Department of Emergency and Critical Care Medicine, St.Marianna University School of Medicine, Kawasaki, Japan; Japanese Red Cross Medical Center, Tokyo, Japan; Matsumoto City Hospital, Matsumoto, Japan; Department of Emergency and Critical Care Medicine, Faculty of Medicine, Fukuoka University, Fukuoka, Japan; Department of Infection Control, Fukuoka University Hospital, Fukuoka, Japan; Department of Rheumatology, National Hospital Organization Hokkaido Medical Center, Sapporo, Japan; Department of Respiratory Medicine, National Hospital Organization Hokkaido Medical Center, Sapporo, Japan; Department of Emergency and Critical Care Medicine, National Hospital Organization Hokkaido Medical Center, Sapporo, Japan; NHO Kanazawa Medical Center, Kanazawa, Japan; Nihon University School of Medicine, Department of Internal Medicine, Division of Respiratory Medicine, Tokyo, Japan; Musashino Red Cross Hospital, Musashino, Japan; Division of Respiratory Medicine, Social Welfare Organization Saiseikai Imperial Gift Foundation, Inc., Saiseikai Kumamoto Hospital, Kumamoto, Japan; Department of Respiratory Medicine, Nagoya University Graduate School of Medicine, Nagoya, Japan; Fukuoka Tokushukai Hospital, Department of Internal Medicine, Kasuga, Japan; Fukuoka Tokushukai Hospital, Respiratory Medicine, Kasuga, Japan; Department of Endocrinology, Hematology and Gerontology, Chiba University Graduate School of Medicine, Chiba, Japan; Department of Emergency and Critical Care Medicine, Chiba University Graduate School of Medicine, Chiba, Japan; National Hospital Organization Kumamoto Medical Center, Kumamoto, Japan; Division of Infectious Diseases and Respiratory Medicine, Department of Internal Medicine, National Defense Medical College, Tokorozawa, Japan; Sapporo City General Hospital, Sapporo, Japan; Department of Internal Medicine, Tokyo Saiseikai Central Hospital, Tokyo, Japan; Department of Pulmonary Medicine, Tokyo Saiseikai Central Hospital, Tokyo, Japan; Department of Respiratory Medicine, Fujisawa City Hospital, Fujisawa, Japan; Uji-Tokushukai Medical Center, Uji, Japan; Department of Infectious Disease and Clinical Research Institute, National Hospital Organization Kyushu Medical Center, Fukuoka Japan; Department of Respirology, National Hospital Organization Kyushu Medical Center, Fukuoka, Japan; Division of Respirology, Rheumatology, and Neurology, Department of Internal Medicine, Kurume University School of Medicine, Kurume, Japan; Department of Infectious Disease, National Hospital Organization Kyushu Medical Center, Fukuoka Japan; Ome Municipal General Hospital, Ome, Japan; Research Institute for Diseases of the Chest, Graduate School of Medical Sciences, Kyushu University, Fukuoka, Japan; Department of Medicine and Biosystemic Science, Kyushu University Graduate School of Medical Sciences, Fukuoka, Japan; Daini Osaka Police Hospital, Osaka, Japan; Department of Emergency and Critical Care Medicine, Faculty of Medicine, University of Tsukuba, Tsukuba, Japan; Department of Hematology, Faculty of Medicine, University of Tsukuba, Tsukuba, Japan; Department of Nephrology, Faculty of Medicine, University of Tsukuba, Tsukuba, Japan; Department of Cardiovascular Surgery, Faculty of Medicine, University of Tsukuba, Tsukuba, Japan; Division of Pulmonary Medicine, Department of Medicine, Tokai University School of Medicine, Isehara, Japan; Department of Anesthesiology and Intensive Care Medicine, Kyoto Prefectural University of Medicine, Kyoto, Japan; Department of Infection Control and Laboratory Medicine, Kyoto Prefectural University of Medicine, Kyoto, Japan; Department of Respiratory Internal Medicine, St Marianna University School of Medicine, Yokohama-City Seibu Hospital, Yokohama, Japan; KINSHUKAI Hanwa The Second Hospital, Osaka, Japan; Gifu University School of Medicine Graduate School of Medicine, Emergency and Disaster Medicine, Gifu, Japan; Department of Respiratory Medicine, Tokyo Medical University Hospital, Tokyo, Japan; JA Toride medical hospital, Toride, Japan; Okayama Rosai Hospital, Okayama, Japan; Himeji St. Mary’s Hospital, Himeji, Japan; Emergency & Critical Care, Niigata University, Niigata, Japan; Emergency & Critical Care Center, National Hospital Organization Kyoto Medical Center, Kyoto, Japan; National Hospital Organization Tokyo Hospital Hospital, Kiyose, Japan; Fujioka General Hospital, Fujioka, Japan; Department of General Medicine, School of Medicine, International University of Health and Welfare Shioya Hospital, Ohtawara, Japan; Department of Pharmacology, School of Pharmacy, International University of Health and Welfare Shioya Hospital, Ohtawara, Japan; Department of Respiratory Medicine, International University of Health and Welfare Shioya Hospital, Ohtawara, Japan; Department of Clinical Laboratory, International University of Health and Welfare Shioya Hospital, Ohtawara, Japan; Department of Cardiology, Pulmonology, and Nephrology, Yamagata University Faculty of Medicine, Yamagata, Japan; Division of Respiratory Medicine and Allergology, Department of Medicine, School of Medicine, Showa University, Tokyo, Japan; Department of Pulmonary Medicine, Fukushima Medical University, Fukushima, Japan; Kansai Electric Power Hospital, Osaka, Japan; Division of Infectious Diseases, Kumamoto City Hospital, Kumamoto, Japan; Department of Respiratory Medicine, Kumamoto City Hospital, Kumamoto, Japan; Department of Emergency and Critical Care Medicine, Tokyo Metropolitan Police Hospital, Tokyo, Japan; Department of Respiratory Medicine, Gunma University Graduate School of Medicine, Maebashi, Japan; National hospital organization Saitama Hospital, Wako, Japan; Tokyo Medical University Ibaraki Medical Center, Inashiki, Japan; Department of Internal Medicine, Kiryu Kosei General Hospital, Kiryu, Japan; Department of Pulmonary Medicine and Oncology, Graduate School of Medicine, Nippon Medical School, Tokyo, Japan; Genome Medical Science Project (Toyama), National Center for Global Health and Medicine, Tokyo, Japan; Department of Biomolecular Engineering, Graduate School of Tokyo Institute of Technology, Tokyo, Japan; School of Veterinary Medicine, Kitasato University, Towada, Japan; Laboratory of Viral Infection I, Department of Infection Control and Immunology, Ōmura Satoshi Memorial Institute & Graduate School of Infection Control Sciences, Kitasato University, Tokyo, Japan; Department of Insured Medical Care Management, Tokyo Medical and Dental University Hospital of Medicine, Tokyo, Japan; Department of Immunopathology, Immunology Frontier Research Center (WPI-IFReC), Osaka University, Suita, Japan; Center for Infectious Disease Education and Research (CiDER), Osaka University, Suita, Japan; Department of Organoid Medicine, Keio University School of Medicine, Tokyo, Japan; Medical Innovation Promotion Center, Tokyo Medical and Dental University, Tokyo, Japan; Department of Surgery, Keio University School of Medicine, Tokyo, Japan; Institute of Research, Tokyo Medical and Dental University, Tokyo, Japan; Institute for the Advanced Study of Human Biology (WPI-ASHBi), Kyoto University, Kyoto, Japan; Department of Medicine, Center for Hematology and Regenerative Medicine, Karolinska Institute, Stockholm, Sweden; Laboratory of Statistical Immunology, Immunology Frontier Research Center (WPI-IFReC), Osaka University, Suita, Japan

## Abstract

To elucidate the host genetic loci affecting severity of SARS-CoV-2 infection, or Coronavirus disease 2019 (COVID-19), is an emerging issue in the face of the current devastating pandemic. Here, we report a genome-wide association study (GWAS) of COVID-19 in a Japanese population led by the Japan COVID-19 Task Force, as one of the initial discovery GWAS studies performed on a non-European population. Enrolling a total of 2,393 cases and 3,289 controls, we not only replicated previously reported COVID-19 risk variants (e.g., *LZTFL1, FOXP4, ABO*, and *IFNAR2*), but also found a variant on 5q35 (rs60200309-A at *DOCK2*) that was associated with severe COVID-19 in younger (<65 years of age) patients with a genome-wide significant p-value of 1.2 × 10^-8^ (odds ratio = 2.01, 95% confidence interval = 1.58-2.55). This risk allele was prevalent in East Asians, including Japanese (minor allele frequency [MAF] = 0.097), but rarely found in Europeans. Cross-population Mendelian randomization analysis made a causal inference of a number of complex human traits on COVID-19. In particular, obesity had a significant impact on severe COVID-19. The presence of the population-specific risk allele underscores the need of non-European studies of COVID-19 host genetics.

## Introduction

Coronavirus disease 2019 (COVID-19) caused by severe acute respiratory syndrome coronavirus 2 (SARS-CoV-2) has been a serious global public health issue^1^. Although promising vaccines have recently become available, the emergence of SARS-CoV-2 variants may delay the end of this pandemic^2^. One of the clinical characteristics of COVID-19 is its diverse clinical presentation, ranging from asymptomatic infection to fatal respiratory/multi-organ failure. For example, elderly peoples are known to be at high risk for severe diseases, particularly those who have multiple complications, such as obesity, hypertension, diabetes, and chronic renal failures^3–6^. The risk for severe disease and mortality may differ depending on populations; Asians, for instance, may have less severe clinical presentation than Europeans^7^. However, practical explanations on this paradox have been elusive.

Human genetic backgrounds influence susceptibility to and/or severity of infectious diseases. Genome-wide association studies (GWAS) have identified a number of genetic loci associated with infectious diseases, including class I human leukocyte antigen (HLA) and *CCR5* implicated in HIV infection, 18q11 in tuberculosis, *HBB* in malaria infection, and 16p21 in *Mycobacterium avium* complex (MAC)^8–11^. Thus, human host genetics is expected to define the susceptibility to COVID-19 and explain the populational differences in disease mortality and morbidity. The Severe Covid-19 GWAS Group reported that a variant at *LZTFL1* on 3p21 confers a risk for severe COVID-19 in Europeans with an odds ratio (OR) of as high as 2.0^12^. The risk variant at 3p21, known to be inherited from the archaic hominin-derived genome sequences of Neanderthals^13^, is also associated with various clinical manifestations of COVID-19 such as severe respiratory failure and venous thromboembolism^14^. Of interests, this variants demonstrated globally heterogeneous allele frequency spectra: highest in South Asians (>40%), moderate in Europeans (≃10%), but rarely present among East Asians^13^. Despite its dominant impact in Europeans, this region does not explain COVID-19 severities among East Asians.

Further GWAS and whole genome sequencing (WGS) have identified host susceptibility genes. In addition to the *ABO* locus^12^, *OAS1/3/2, TYK2, DPP9*, and *IFNAR2* have been identified^15,16^. WGS revealed rare loss-of-function (LoF) mutations in *TLR7* and type I IFN-related genes in severe COVID-19 patients^17^. In particular, the international consortium COVID-19 Human Genome Initiatives (HGI) has been conducting a large-scale meta-GWAS^15,16^. However, the vast majority of the existing genomic studies were performed on European populations. Considering the global diversity of COVID-19 severity, COVID-19 host genetics analysis in non-European populations should provide novel insights.

The Japan COVID-19 Task Force was established in early 2020 as a nation-wide multicenter consortium to overcome the COVID-19 pandemic in Japan (https://www.covid19-taskforce.jp/en/home/; **Figure 1**). This collaborative research networks are composed of >100 hospitals throughout Japan led by core academic institutes (**Supplementary Table 1**). Since the beginning of the pandemic, the Japan COVID-19 Task Force has longitudinally collected DNA, RNA, and plasma from >3,400 COVID-19 cases along with detailed clinical information (as of April 2021). The collected bioresources have been invested for multi-omics analysis towards open science for the community. In this study, we report the result of an initial large-scale discovery GWAS of COVID-19 in a Japanese population with systemic comparisons to that in Europeans to elucidate the global host genetics landscape of COVID-19.

**Table 1.** Association of the DOCK2 variant with COVID-19 risk in the Japanese population.

| rsID | Chr:position<br>(b37) | Allele | Gene | Age | Phenotype | No. subjects |  | Risk allele freq. (A) |  | OR (95%CI) | P |
| --- | --- | --- | --- | --- | --- | --- | --- | --- | --- | --- | --- |
|  |  |  |  |  |  | Cases | Controls | Cases | Controls |  |  |
| rs60200309 | 5:169519612 | G/A | DOCK2 | All age | COVID-19<br>vs control | 2,393 | 3,289 | 0.119 | 0.101 | 1.24 (1.09-1.41) | 0.0011 |
|  |  |  |  |  | Severe COVID-19<br>vs control | 990 | 3,289 | 0.129 | 0.101 | 1.39 (1.16-1.66) | 3.1×10<sup>-4</sup> |
|  |  |  |  | Age < 65 | COVID-19<br>vs control | 1,484 | 2,377 | 0.123 | 0.099 | 1.32 (1.13-1.55) | 5.1×10<sup>-4</sup> |
|  |  |  |  |  | Severe COVID-19<br>vs control | 440 | 2,377 | 0.159 | 0.099 | 2.01 (1.58-2.55) | 1.2×10<sup>-8</sup> |

**Figure 1.**
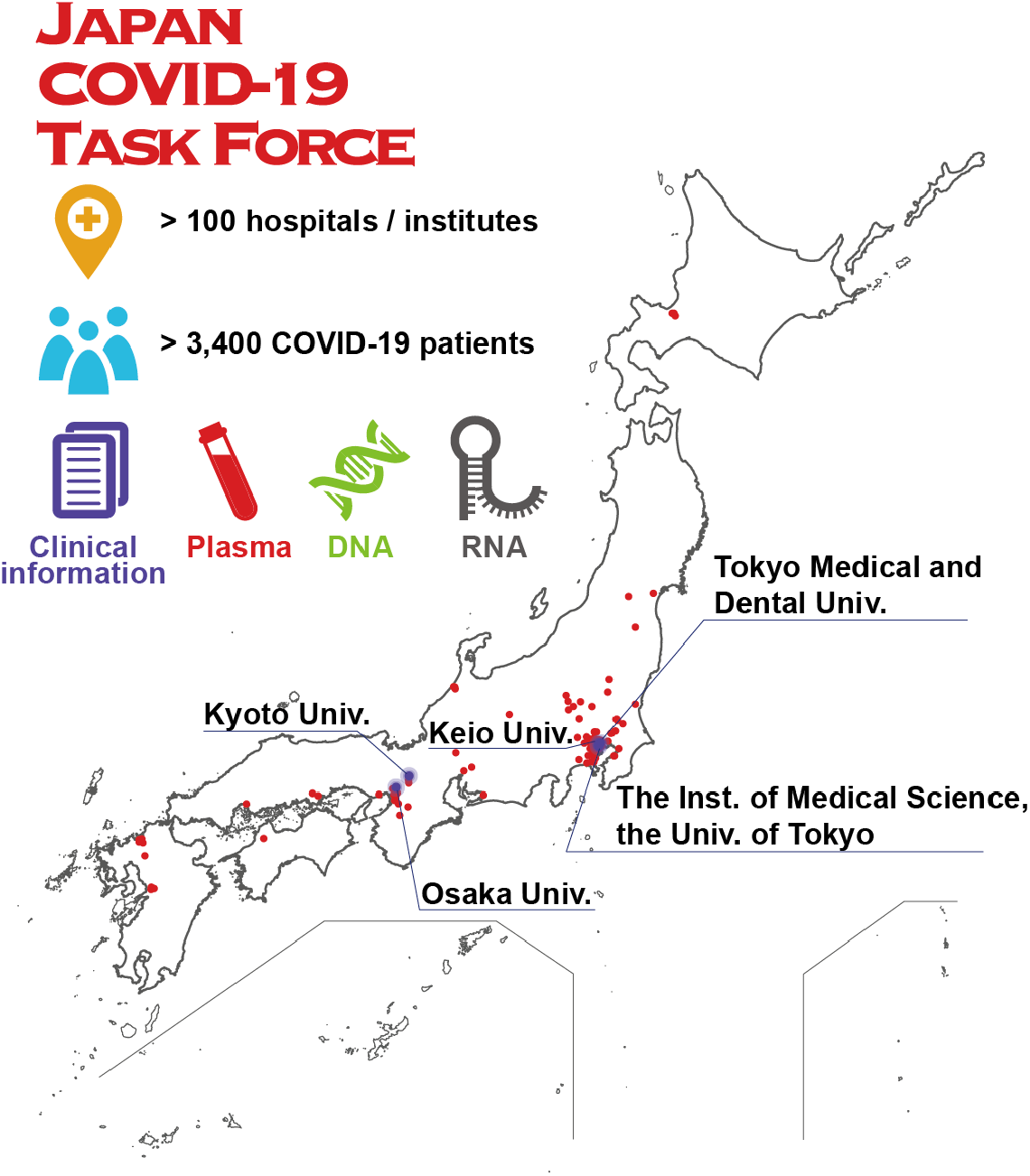
Japan COVID-19 Task Force. Japan COVID-19 Task Force is a nation-wide consortium to overcome COVID-19 pandemic in Japan, which was established in early 2020. Japan COVID-19 Task Force consists of >100 hospitals (red dots) led by core academic institutes (blue labels), and collected DNA, RNA, and plasma from >3,400 COVID-19 cases along with detailed clinical information.

## Results

### Overview of the study participants

In this study, we enrolled unrelated 2,393 patients with COVID-19 who required hospitalization from April 2020 to January 2021, from >100 hospitals participating in the Japan COVID-19 Task Force. COVID-19 diagnoses of all cases were confirmed by physicians of each affiliated hospital based on clinical manifestations and a positive PCR test result. As for the control, we enrolled unrelated 3,289 subjects ahead of the COVID-19 pandemic who represent a general Japanese population. All the participants were confirmed to be of Japanese origin on the basis of a principal component analysis (**Supplementary Figure 1**). Detailed characteristics of the participants are described in **Supplementary Table 2**.

Of the 2,393 COVID-19 cases, 990 ultimately had severe infection as defined by oxygen support, artificial respiration, and/or intensive-care unit hospitalization), while 1,391 cases had non-severe diseases. Severity information was not available for the remaining 12. As reported previously^3,18^, the severe COVID-19 cases were relatively more aged (65.3 ± 13.9 years [mean ± SD]) and a higher proportion of males (73.9%), compared with non-severe cases (49.3 ± 19.2 years and 57.2 of males, respectively).

### GWAS of COVID-19 in Japanese identified a population-specific risk variant at *DOCK2*

We conducted a GWAS of COVID-19 in a Japanese population. After applying stringent quality control (QC) filters and genome-wide genotype imputation using a population-specific reference panel of Japanese^19–21^, we obtained 13,485,123 variants with minor allele frequency (MAF) ≥ 0.001 and imputation score (Rsq) ≥ 0.5 (13,116,003, 368,566 and 554 variants for autosomal, X-chromosomal, and mitochondrial variants, respectively). As illustrated in the follow-up analysis stratified by deep clinical information at the *LZTFL1* locus, several COVID-19 risk variants are expected to confer relatively larger effects in severe and younger cases than in mild (and self-reported) cases or elder cases^14^. This suggests that COVID-19 GWAS likely to have a higher statistical power when focusing on severe and younger cases^12,16,22^. We thus separately conducted stratified GWAS of severe COVID-19 cases (*n*_Case_ = 990), younger cases (age < 65, *n*_Case_ = 1,484), and their combinations (*n*_Case_ = 440), as well as all the cases (*n*_Case_ = 2,393), in comparisons with the controls. We selected the age of 65 as a threshold, since ages ≥65 years is defined as a aggravation risk factor in the clinical management guide of patients with COVID-19 in Japan^4^. We did not observe inflation of GWAS test statistics (λ_GC_ < 1.007; **Supplementary Figure 2**), suggesting no evidence of population stratification, as well as potential biases, in our GWAS.

**Figure 2.**
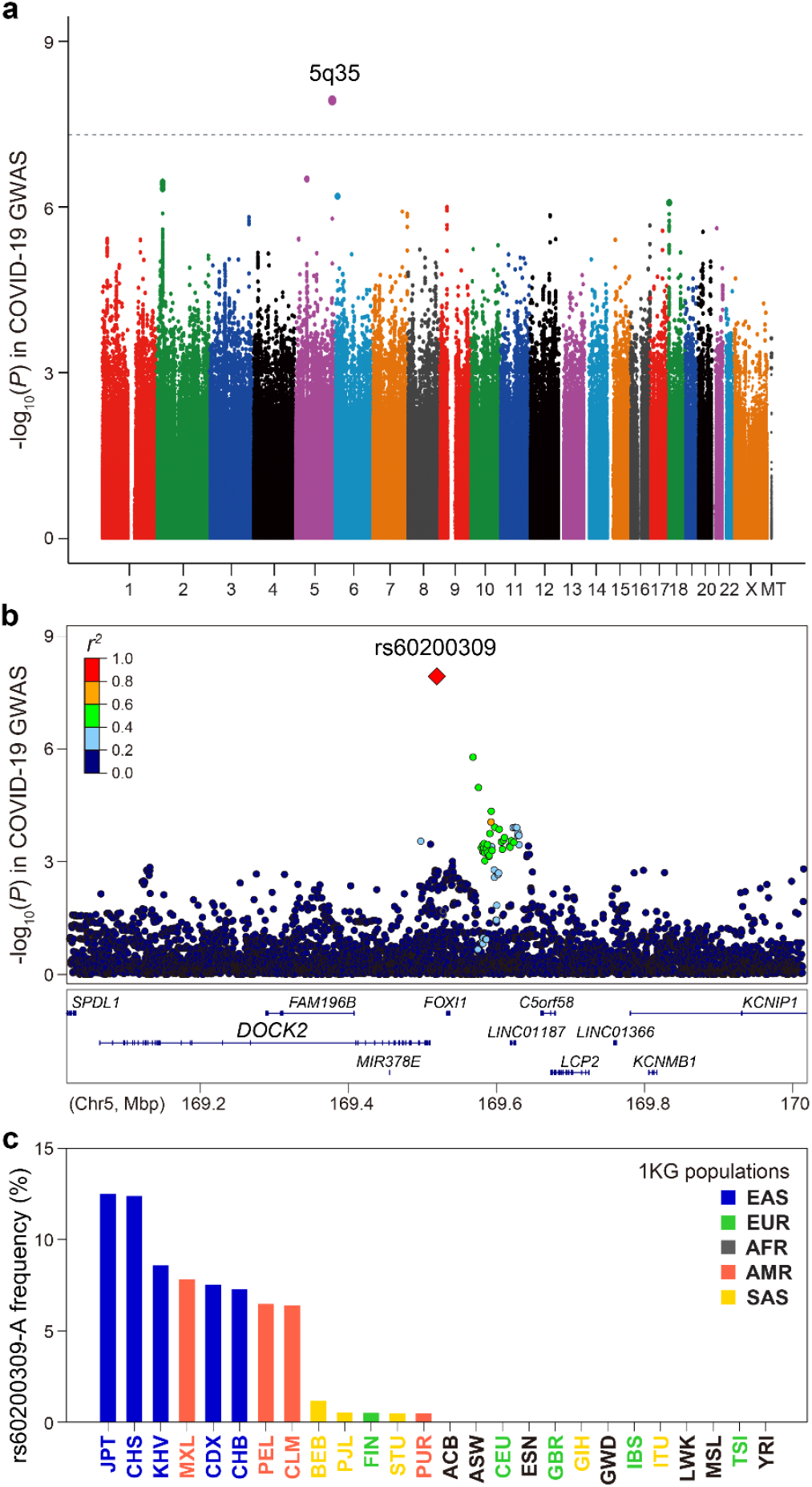
Severe and younger COVID-19 GWAS in the Japanese population. (**a**) A Manhattan plot of the severe and younger COVID-19 GWAS (440 cases and 2,377 controls). A dotted line represents the genome-wide significance threshold of *P* < 5.0 × 10^-8^. Manhattan and quantile-quantile plots of all GWAS results are in **Supplementary Figure 2**. (**b**) A regional association plot at the *DOCK2* locus. Dots represent SNPs with colors according to linkage disequilibrium (*r*^*2*^) with the lead SNP of rs60200309. (**c**) Allele frequency spectra of the rs60200309-A allele in the 1000 Genomes Project Phase3v5 database.

GWAS between all COVID-19 cases vs. controls yielded no positive signals satisfying a genome-wide significance threshold of *P* < 5.0 × 10^-8^ (**Supplementary Figure 2**)^23^. By contrast, when the comparison was made between younger cases with sever COVID-19 and respective controls, where the highest prior probability of discovery of a positive association was expected, we identified a genetic locus on 5q35 that satisfied genome-wide significance (*P* = 1.2 × 10^-8^ at rs60200309; **Figure 2a**). The A allele of the lead SNP (rs60200309) that was located at an intergenic region downstream of the *DOCK2* gene was associated with an inflated risk for severe COVID-19 infection with an OR of 2.01 (95% confidence interval [95%CI] = 1.58-2.55, *P* = 1.2 × 10^-8^; **Table 1** and **Figure 2b**). The risk rs60200309-A allele was also associated with a significantly increased risk of COVID-19 in other comparisons including all COVID-19 cases and controls regardless of severity (OR = 1.24, 95%CI = 1.09-1.41, *P* = 0.0011; **Supplementary Table 3**), and within-case severity analysis (i.e., severe vs non-severe cases; OR = 1.27, 95%CI = 1.03-1.57, *P* = 0.028 for all ages; OR = 1.90, 95%CI = 1.43-2.52, *P* = 1.1 × 10^-5^ for ages <65). To evaluate the effect of the time of enrollment, we conducted a stratified analysis according to the time of enrollment for younger cases with severe COVID-19. The rs60200309-A allele consistently showed an inflated risk throughout the recruitment period (OR = 2.38, 1.79, 1.92 in the stratified analysis according to the trisected recruitment periods of April 2020 – July 2020, August 2020 – October 2020, and November 2020 – January 2021, respectively; OR = 2.00 and *P* = 2.0 × 10^-8^ in the all-period meta-analysis; **Supplementary Table 3**). This allele, however, did not seem to confer any significant COVID-19 risk in elder cases (*P* > 0.069). These results suggest a susceptibility of patients with the rs60200309-A allele to severe COVID-19 in the Japanese population, particularly in younger cases with severe COVID-19.

We then looked up COVID-19 risk of the *DOCK2* variant in different ancestries (3,138 hospitalized COVID-19 cases vs 891,375 controls from the pan-ancestry meta-analysis available at https://rgc-covid19.regeneron.com/)^24,25^. We observed the same directional effect with a marginal association signal (OR = 1.73, 95%CI = 0.95-3.15, *P* = 0.072, MAF_Case_ = 0.0025, MAF_Control_ = 0.0008; **Supplementary Table 4**). Meta-analysis of the Japanese discovery GWAS and the replication study from the pan-ancestry study yielded a genome-wide significant association showing an OR = 1.97 (95%CI = 1.57-2.46, *P* = 1.2 × 10^-9^; **Supplementary Table 5**). We note that rs60200309 does not exist in the public summary statistics provided by the COVID-19 Host Genetics Initiative (release 5)^16^. Nevertheless, given the low allele frequencies of the relevant *DOCK2* allele in non-East Asian populations, further population-specific and cross-population replication studies are warranted.

### Population-specific features of the *DOCK2* risk allele associated with COVID-19

Of interest, the risk allele at *DOCK2* (rs60200309-A) identified in this study was common in East Asians (= 0.097) with the highest frequency in Japanese (= 0.125), less frequent in native Americans (= 0.049), but very rare in Europeans, African, and south Asians (< 0.005; from 1000 Genomes Project Phase3v5 database; **Figure 2c**). When we referred to the results of WGS-based natural selection screening in Japanese^19^, the rs60200309-A allele marginally positively selected in Japanese (*P*_SDS_ = 0.051). It is suggested that the rs60200309-A rapidly increased its frequency among the Japanese population during the past several thousand years. These population-specific features of the *DOCK2* variant partly explain the reason why it was not identified in the previous European COVID-19 GWAS studies despite their larger sample sizes, and should provide a rationale for further accelerating COVID-19 host genetics researches on non-European populations.

### Cross-population comparisons of the COVID-19-associated variants

We then conducted cross-population comparisons of allele frequency spectra and genetic risk of the previously reported COVID-19-associated variants^12,16,22^. Of the 11 associated variants evaluated in our Japanese GWAS, we replicated the associations with 7 variants (*P* < 0.05 in any 4 phenotypes of the case-control GWAS; *LZTFL1, FOXP4, TMEM65, ABO, TAC4, DPP9*, and *IFNAR2*; **Figure 3** and **Supplementary Table 6**). For all nominally associated signals (*P* < 0.05), we observed same directional effects of the alleles as in Europeans. ORs for severe and younger COVID-19 cases were highest among the phenotype patterns in six of the 7 loci, confirming our strategy that focusing on such cases should efficiently highlight the host genetic risk of COVID-19.

**Figure 3.**
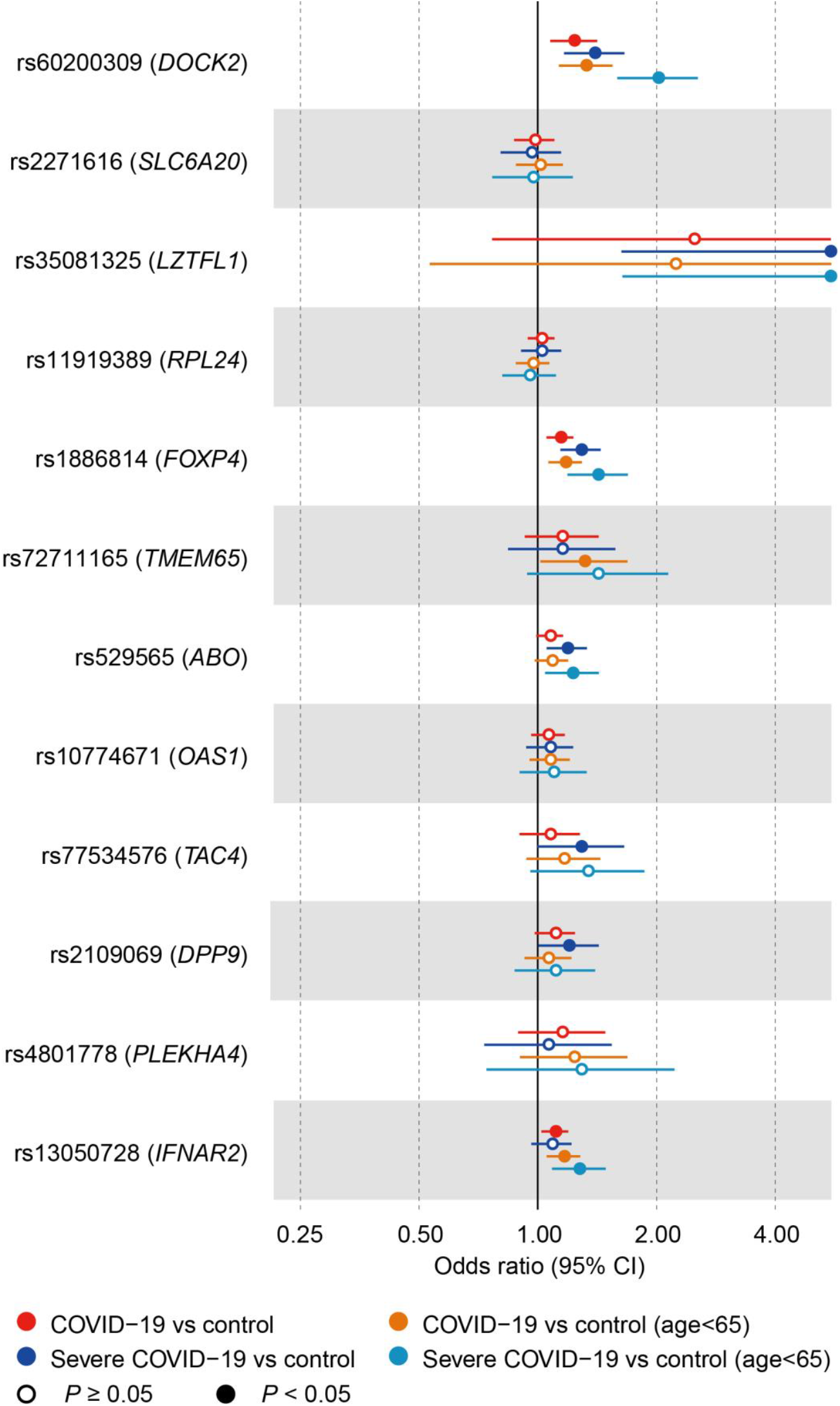
Forest plots of the risk of COVID-19-associated variants in Japanese. Odds ratios of the COVID-19-associated variants in the Japanese population are indicated.

The most significant replication was observed at the *FOXP4* locus, where the risk allele was known to be more prevalent in East Asians than in Europeans, and expected to have a higher power to be detected in East Asians^16^ (OR = 1.29, 95%CI = 1.13-1.46, *P* = 9.1 × 10^-5^ for the severe COVID-19 cases). Regarding the strongest risk variant in Europeans at *LZTFL1* (rs35081325), we replicated the associations despite its rare frequency in Japanese (= 0.0013 in controls) with the highest risk in the severe and younger COVID-19 cases (OR = 11.8, 95%CI = 1.64-85.5, *P* = 0.014). These observations propose shared host genetics backgrounds of COVID-19 across populations.

### Relatively less dominant impact of the HLA gene variants on COVID-19 risk

Given their critical impact on immune responses and contribution to host genetics of various infectious diseases,^26,27^ HLA gene variants have been investigated for their possible role in the response to COVID-19 infection with controversial discussions^28,29^. To address this issue, we applied *in silico* imputation of both classical and non-classical HLA variants using the HLA reference panel of Japanese (*n* = 1,118)^30,31^. After imputing the HLA variants, we did not observe association signals satisfying neither of the genome-wide significance (*P* < 5.0 × 10^-8^) or HLA-wide significance thresholds (*P* < 0.05/2,482 variants = 2.0 × 10^-5^; **Supplementary Figure 3** and **Supplementary Table 7**). Most significant HLA variant associations in each of the case-control phenotypes were followings: all cases (HLA-DQα1 Glu175, OR = 0.84, 95%CI = 0.77-0.92, *P* = 2.9 × 10^-4^), severe cases (HLA-DRβ1 amino acid position 38, *P* = 6.7 × 10^-4^), younger cases (HLA-DRβ1 amino acid positions 96 and 104, *P* = 8.3 × 10^-5^), and severe and younger cases (HLA-C*03:04, OR = 1.48, 95%CI = 1.16-1.90, *P* = 0.0020). While further accumulation of sample sizes is warranted, the current results may not support a major impact of the HLA variants on host genetics in Japanese despite previous associations with other infectious diseases.

### Link between ABO blood type and COVID-19 risk

ABO blood types are defined by the variants on the coding region of the *ABO* gene on 9q34^32^, which are pleiotropic on various complex human traits including infectious diseases (e.g., malaria resistance of blood type O^33^). Motivated by replicated COVID-19 risk of the *ABO* locus in Japanese, we conducted ABO blood type-based risk analysis.^34^

Among the four major ABO blood types (A, B, AB, and O with 39.0%, 21.8%, 9.5%, and 29.7% in our Japanese GWAS, respectively), the O blood type was consistently associated with a protective effect on COVID-19 in case-control phenotypes (*P* < 0.05), most evidently in severe and younger cases (OR = 0.73, 95%CI = 0.56-0.93, *P* = 0.014; **Figure 4** and **Supplementary Table 8**), as reported previously^12^. we found increased risk of the AB blood type, especially in severe cases (OR = 1.41, 95%CI = 1.10-1.81, *P* = 0.0065 for all ages, and OR = 1.40, 95%CI = 1.00-1.94, *P* = 0.048 for age < 65, in comparison with the other blood types). Increased severity risk of the AB blood type was also significant when compared with the A or B blood types (OR > 1.34, *P* < 0.041 for all ages). To our knowledge, this is the initial study to report severe COVID-19 risk of the AB blood type. The ABO blood type distributions are heterogeneous among worldwide populations, and Japanese is the one with the highest AB blood type frequency^35^, which might have provided statistical power to detect its risk on severe COVID-19 in our study.

**Figure 4.**
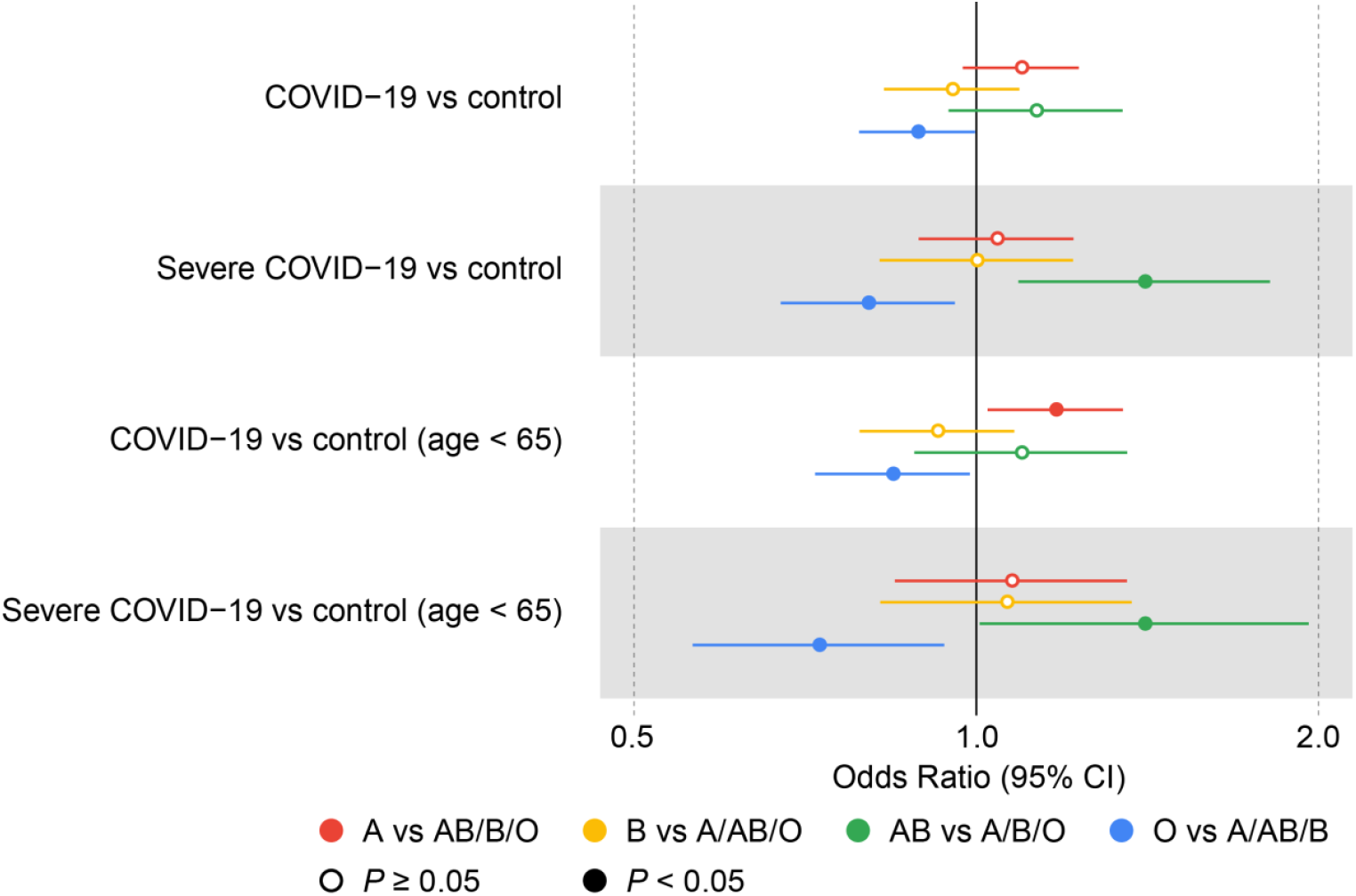
Forest plots of the ABO blood type associations with COVID-19 in Japanese. Odds ratios of the ABO blood types in the Japanese population are indicated.

### Causal inference on COVID-19 by cross-population Mendelian randomization

COVID-19 pandemic has exposed global populations to an emergent health risk. To predict individuals’ risk on COVID-19-related outcomes, elucidation of the medical conditions that can affect COVID-19 susceptibility is warranted. While medical record-based epidemiological studies assessing comorbidity have identified multiple risk factors, there remains various clinical status where causal inference on COVID-19 is controversial^3^.

To make a causal inference, we applied cross-population two-sample Mendelian randomization (MR) analysis. Two-sample MR utilizes GWAS summary statistics to infer causality between correlated phenotypes.^36^ Further, cross-population MR analysis could provide robust evidence of causality^37^. As exposure phenotypes, we selected a series of clinical states and diseases where increased or decreased comorbidity with COVID-19 have been discussed: metabolic traits (obesity [body mass index; BMI] and type 2 diabetes [T2D]), chronic respiratory diseases and related phenotypes (cigarettes per day [CPD] and asthma), blood pressure, renal function (estimated glomerular filtration rate [eGFR]), serum uric acids (UA) and gout, and rheumatic diseases (rheumatoid arthritis [RA] and systemic lupus erythematosus [SLE])^3–6,18,38,39^. Lists of the GWAS studies of the exposure phenotypes are in **Supplementary Table 9**.

In the Japanese population, MR results were contrastive between the severe COVID-19 cases and all COVID-19 cases (**Figure 5** and **Supplementary Table 10**). As for the severe COVID-19 cases, a causal effect was demonstrated only for obesity (*P* = 0.0067 and 0.0074 for all age and age < 65, respectively). By contrast, we observed causal effects of asthma (*P* = 0.0061 and 0.018 for all age and age < 65, respectively), UA (*P* = 0.019 for age < 65), and gout (*P* = 0.0048 and 0.0027 for all age and age < 65, respectively), while SLE (*P* = 0.0014 for all age) showed a protective effect.

**Figure 5.**
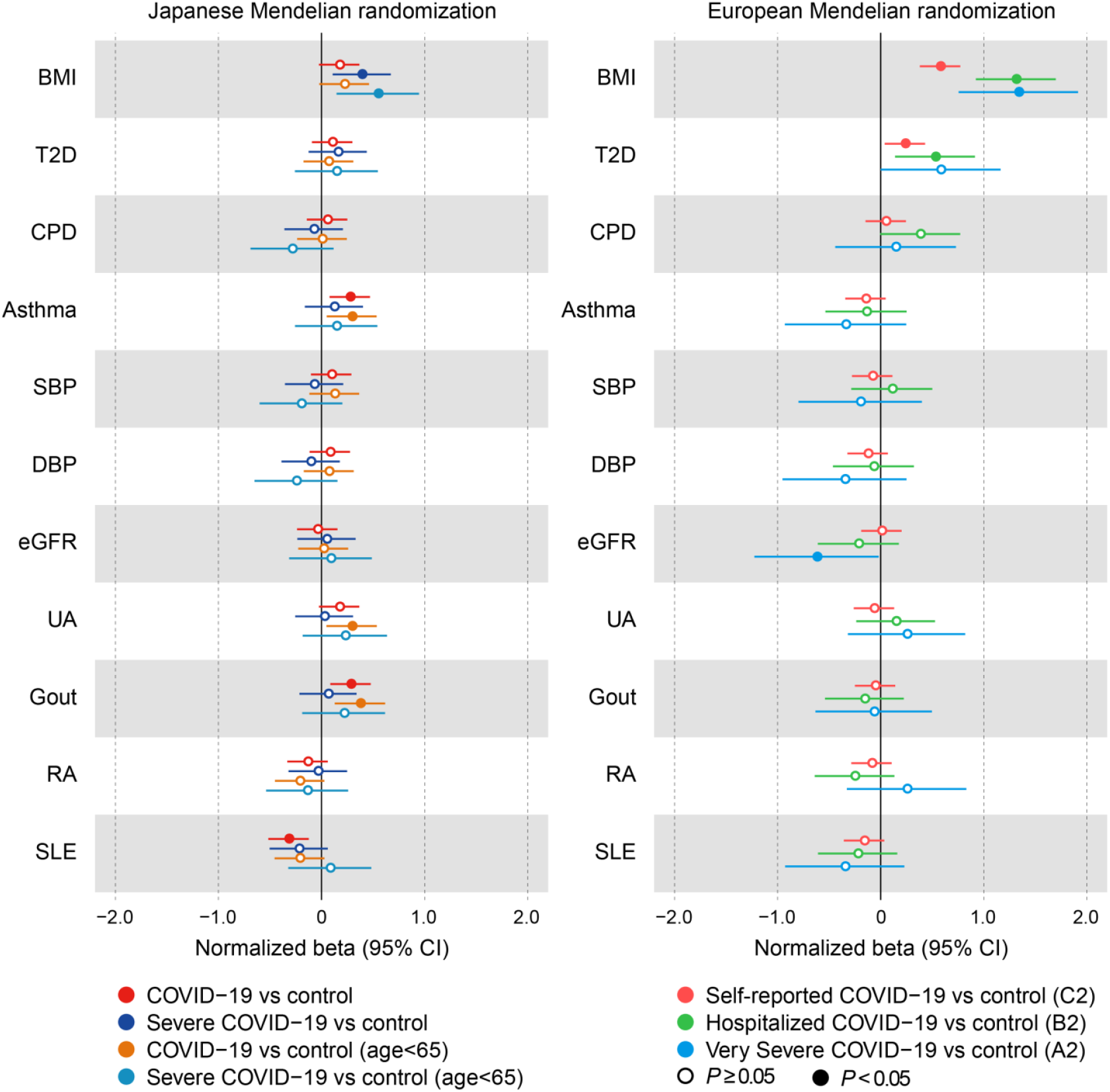
Cross-population Mendelian randomization analysis of the COVID-19 GWAS. Forest plots of the Mendelian randomization (MR) analysis results of causal inference on the COVID-19 GWAS in Japanese (left panel) and Europeans (right panel). Since effect sizes (= beta) of MR are not scalable among the phenotypes and populations, normalized beta is indicated. For each phenotype and population, the standard error for the COVID-19 GWAS with the largest sample size (i.e., “COVID-19 vs control” for Japanese and “Self-reported COVID-19 vs control (C2)” for Europeans) was set to be 0.1. Abbreviations of the exposure phenotypes and the detailed MR results are in **Supplementary Table 10**.

We then looked up the MR results in Europeans by using publicly released GWAS summary statistics of COVID-19 Host Genetics Initiative (release 5)^16^. We observed significant causal inferences of obesity consistent with those in Japanese. The causal effect of obesity was observed for self-reported, hospitalized, and severe COVID-19 in Europeans (*P* = 8.5 × 10^-9^, 3.2 × 10^-11^, and 6.2 × 10^-6^, respectively) as previously reported^40^, while effect sizes were twice as high in hospitalized and severe COVID-19 (*β* > 0.398) when compared with self-reported COVID-19 (*β* = 0.175). Obesity is one of the major risk factors for COVID-19 severity and critical outcomes^3–5^, and our cross-population MR analysis provided evidence of causality on this link. Causal inference of decreased renal function (*P* = 0.043 for severe COVID-19) and T2D (*P* = 0.019 for self-reported COVID-19 and *P* = 0.0078 for hospitalized COVID-19) was also observed in Europeans.

Our cross-population MR analysis provided several phenotypes with significant MR results observed only in Japanese (i.e., risk of asthma, UA, and gout, and protective role of SLE). This suggests existence of populational heterogeneity in the impacts of causal links from the baseline clinical manifestations to COVID-19 susceptibility. Hyperuricemia is reported as one of the major risk factors of severe COVID-19 in Japan^18^, which is consistent with a Japanese-specific causal inference of UA and gout in the Japanese MR analysis. There exist controversial discussions on the risk of SLE patients on COVID-19 infection^38,39^. Our results suggest a possibility that genetically-determined susceptibility to SLE, and its underlying immunophenotypes, could make patients protective against COVID-19 infection.

## Discussion

In this study, we reported GWAS of COVID-19 in a Japanese population led by the Japan COVID-19 Task Force, a nation-wide consortium to battle against the COVID-19 pandemic. This is one of the initial and largest COVID-19 host genetics studies in non-European populations to date. Our study highlighted multiple genetic variants associated with the COVID-19 risk shared across populations such as *LZTFL1, ABO*, and *FOXP4*, as well as the identification of a population-specific risk variant at the *DOCK2* locus. Stratified analysis of these susceptibility loci supported the expectation that host genetics of COVID-19 should be enhanced when focusing the analysis on younger cases with severe COVID-19. Rather unexpectedly, contribution of HLA variants to COVID-19 host susceptibility, if ever present, was not remarkable, compared with previous findings on other infectious diseases. As for the ABO blood type classification, we newly identified the risk of the AB blood type to severe COVID-19. Finally, cross-population MR analysis disclosed a causal inference of a number of complex human traits on COVID-19, such as an elevated risk of obesity on severe COVID-19. Our results highlight population-specific risk allele host genetic backgrounds, which underscores the need of non-European studies for COVID-19 host genetics.

DOCK2 (dedicator of cytokinesis 2) is a Rac activator involved in chemokine signaling, type I interferon production, and lymphocyte migration^41,42^, where the pathophysiology of COVID-19 have been implicated^43,44^. Multiple COVID-19 risk alleles elucidated by host genetics studies are in this axis (e.g., *LZTFL1-SLC6A20-CCR9, TYK2, IFNAR2*, and *IRF7*)^12,16,22^. Of note, autosomal recessive *DOCK2* deficiency is a Mendelian disorder with combined immunodeficiency and severe invasive infection (OMIM #616433)^45^. *DOCK2* has recently been reported to be suppressed in bronchoalveolar lavage fluid (BALF) cells of COVID-19 patients^46^. Given that LoF caused by an inborn error is a key to fine-map host susceptible genes for infectious diseases^26^, *DOCK2* could be considered as one of the key genes to determine the risk for, as well as potential targets, of COVID-19 therapy and drug discovery. Further functional studies linking the *DOCK2* variant to molecular and clinical phenotypes should be required to elucidate the mechanism by which this *DOCK2* allele confer the risk of severe COVID-19.

While our nation-wide longitudinal efforts have enabled the current COVID-19 GWAS in Japanese, a number of replication studies involving independent populations are required to confirm the current result. In the near future, a growing number of GWAS studies should be conducted regarding COVID-19 host genetics, which through public sharing contribute to guide a global health strategy against the COVID-19 pandemic.

## Methods

### Study participants

All the cases affected with COVID-19 were recruited through Japan COVID-19 Task Force. We enrolled the hospitalized cases diagnosed as COVID-19 by physicians using the clinical manifestation and PCR test results, who were recruited from April 2020 to January 2021 at any of the >100 the affiliated hospitals (**Supplementary Table 1 and 2**). Control subjects were collected as general Japanese populations at Osaka University Graduate School of Medicine and affiliated institutes. Individuals determined to be of non-Japanese origin either of self-reporting or by principal component analysis were excluded as described elsewhere (**Supplementary Figure 1**)^47^. All the participants provided written informed consent as approved by the ethical committees of Keio University School of Medicine, Osaka University Graduate School of Medicine, and affiliated institutes.

### GWAS genotyping and quality control

We performed GWAS genotyping of the 2,520 COVID-19 cases and 3,341 controls using Infinium Asian Screening Array (Illumina, CA, USA). We applied stringent QC filters to the samples (sample call rate < 0.97, excess heterozygosity of genotypes > mean + 3SD, related samples with PI_HAT > 0.175, or outlier samples from East Asian clusters in principal component analysis with 1000 Genomes Project samples), and variants (variant call rate < 0.99, significant call rate differences between cases and controls with *P* < 5.0 × 10^-8^, deviation from Hardy-Weinberg equilibrium with *P* < 1.0 × 10^-6^, or minor allele count < 5), as described elsewhere^48^. Details of the QC for the mitochondrial variants are described elsewhere^21^. After QC, we obtained genotype data of 489,539, 15,161, and 217 autosomal, X-chromosomal, and mitochondrial variants, respectively, for 2,393 COVID-19 cases and 3,289 controls.

### Genome-wide and HLA genotype imputation

We used SHAPEIT4 software (version 4.1.2) for haplotype phasing of autosomal genotype data, and SHAPEIT2 software (v2.r904) for X-chromosomal genotype data. After phasing, we used Minimac4 software (version 1.0.1) for genome-wide genotype imputation. We used the population-specific imputation reference panel of Japanese (*n* = 1,037) combined with 1000 Genomes Project Phase3v5 samples (*n* = 2,504)^19,20^. Imputations of the mitochondrial variants were conducted as described elsewhere^21^, using the population-specific reference panel (*n* = 1,037). We applied post-imputation QC filters of MAF ≥ 0.1% and imputation score (Rsq) > 0.5. We note that the genotypes of the lead variant in the GWAS (rs60200309) were obtained by imputation (Rsq = 0.88). We assessed accuracy by comparing the imputed dosages with WGS data for the part of the controls (*n* = 236), and confirmed high concordance rate of 97.5%.

HLA genotype imputation was performed using DEEP*HLA software (version 1.0), a multitask convolutional deep learning method^31^. We used the population-specific imputation reference panel of Japanese (*n* = 1,118), which included both classical and non-classical HLA gene variants for imputation^30^. Before imputation, we removed the overlapping samples between the GWAS controls and the reference panel (*n* = 649), from the GWAS data side. We imputed HLA alleles (2-digit and 4-digit) and the corresponding HLA amino acid polymorphisms, and applied post-imputation QC filters of MAF ≥ 0.5% and imputation score (*r*^*2*^ in cross-validation) > 0.7.

### Case-control association test

We conducted GWAS of COVID-19 by using logistic regression of the imputed dosages of each of the variants on case-control status, using PLINK2 software (v2.00a3LM AVX2 Intel [6 Jul 2020]). We included sex, age, and the top five principal components as covariates in the regression model. We set the genome-wide association significance threshold of *P* < 5.0 × 10^-8 23^. We obtained the association of the *DOCK2* variant (rs60200309) from the pan-ancestry meta-analysis available at https://rgc-covid19.regeneron.com/^24,25^. We obtained the meta-analysis results of the phenotype of “hospitalized COVID-19 vs COVID-19 negative or COVID-19 status unknown” with the largest case sample size. Meta-analysis of the Japanese discovery GWAS and the pan-ancestry analysis was conducted using an inverse-variance method assuming a fixed-effects model.

As for the imputed HLA variants, we conducted (i) association test of binary HLA markers (2-digit and 4-digit HLA alleles, respectively amino acid residues) and (ii) an omnibus test of each of the HLA amino acid positions, as described elsewhere^30^. Binary maker test was conducted using the same logistic regression model and covariates as in the GWAS. Omnibus test was conducted by a log likelihood ratio test between the null model and the fitted model, followed by a χ^2^ distribution with *m*-1 degree(s) of freedom, where *m* is the number of the residues. *R* statistical software (version 3.6.0) was used for the HLA association test. In addition to the genome-wide significance threshold, we set the HLA-wide significance threshold based on Bonferroni’s correction for the number of the HLA tests (*α* = 0.05).

### Estimation of the ABO blood types and analysis

We estimated the ABO blood types of the GWAS subjects based on the five coding variants at the *ABO* gene (rs8176747, rs8176746, rs8176743, rs7853989, and rs8176719)^32,33^. We phased the haplotypes of these five variants based on the best-guess genotypes obtained by genome-wide imputation, and estimated the ABO blood type as described elsewhere^34^. We could unambiguously determine the ABO blood type of 99.1 % of the subjects.

Blood group-specific ORs were estimated based on comparisons of A vs AB/B/O, B vs A/AB/O, AB vs A/B/O, and O vs A/AB/B. We conducted a logistic regression analysis including age, sex and the top 5 principal components as covariates. *R* statistical software (version 3.6.3) was used for the ABO blood type analysis.

### Cross-population MR analysis

We conducted two-sample MR analysis as described elsewhere^36,37^. As an outcome phenotype, we utilized the GWAS summary statistics of Japanese (current study) and Europeans (release 5 from COVID-19 Host Genetics Initiative^16^). Lists of the Japanese and European GWAS studies used as the exposure phenotypes are in **Supplementary Table 9**. We extracted the independent lead variants with genome-wide significance (or the proxy variants in linkage disequilibrium *r*^*2*^ ≥ 0.8 in the EAS or EUR subjects of the 1000 Genomes Project Phase3v5 databases) from the GWAS results of the exposure phenotypes. We applied the inverse variance weighted (IVW) method using the TwoSampleMR package (version 0.5.5) in R statistical software (version 4.0.2).

## Data Availability

GWAS summary statistics of the study will be publicly available.

## Acknowledgements

We would like to sincerely thank all the participants involved in this study, and all the members of Japan COVID-19 Task Force for their supports. We thank Mr. Johji Kitano, e-Parcel Corporation, and Ascend Corporation for voluntarily supporting Japan COVID-19 Task Force. We thank COVID-19 Host Genetics Initiative for publicly sharing the GWAS summary statistics of COVID-19. This study was supported by AMED (JP20nk0101612, JP20fk0108415, JP21jk0210034, JP21km0405211, and JP21km0405217), JST CREST (JPMJCR20H2), MHLW (20CA2054), Takeda Science Foundation, the Mitsubishi Foundation, and Bioinformatics Initiative of Osaka University Graduate School of Medicine, Osaka University. The super-computing resource was provided by Human Genome Center (the Univ. of Tokyo).

## Author contributions

H.Namkoong, K.Fukunaga, T.Ueno, K.Katayama, M.Ai, A.Kumanogoh, Toshiro.Sato, N.Hasegawa, K.Tokunaga, M.Ishii, R.Koike, Yuko.Kitagawa, A.Kimura, S.Imoto, S.Miyano, Seishi.Ogawa, T.Kanai, and Y.Okada designed the study. H.Namkoong, R.Edahiro, K.Fukunaga, Y.Shirai, K.Sonehara, H.Tanaka, H.Lee, T.Hasegawa, Masahiro.Kanai, Tatsuhiko.Naito, K.Yamamoto, R.Saiki, A.Kimura, S.Imoto, S.Miyano, Seishi.Ogawa, T.Kanai, and Y.Okada conducted experiments or data analysis, and wrote the manuscript. H.Namkoong, R.Edahiro, K.Fukunaga, Y.Shirai, K.Sonehara, H.Tanaka, H.Lee, T.Hasegawa, Masahiro.Kanai, Tatsuhiko.Naito, K.Yamamoto, R.Saiki, Y.Nannya, T.Ueno, K.Katayama, M.Ai, A.Kumanogoh, Toshiro.Sato, N.Hasegawa, K.Tokunaga, M.Ishii, R.Koike, Yuko.Kitagawa, A.Kimura, S.Imoto, S.Miyano, Seishi.Ogawa, T.Kanai, and Y.Okada managed collection of the samples. All authors contributed to sample and clinical data collection.

