## Supplementary Table for "Japan COVID-19 Task Force: a nation-wide consortium to elucidate host genetics of COVID-19 pandemic in Japan"

**Supplementary Figure 1. A principal component analysis plot of the GWAS participants**

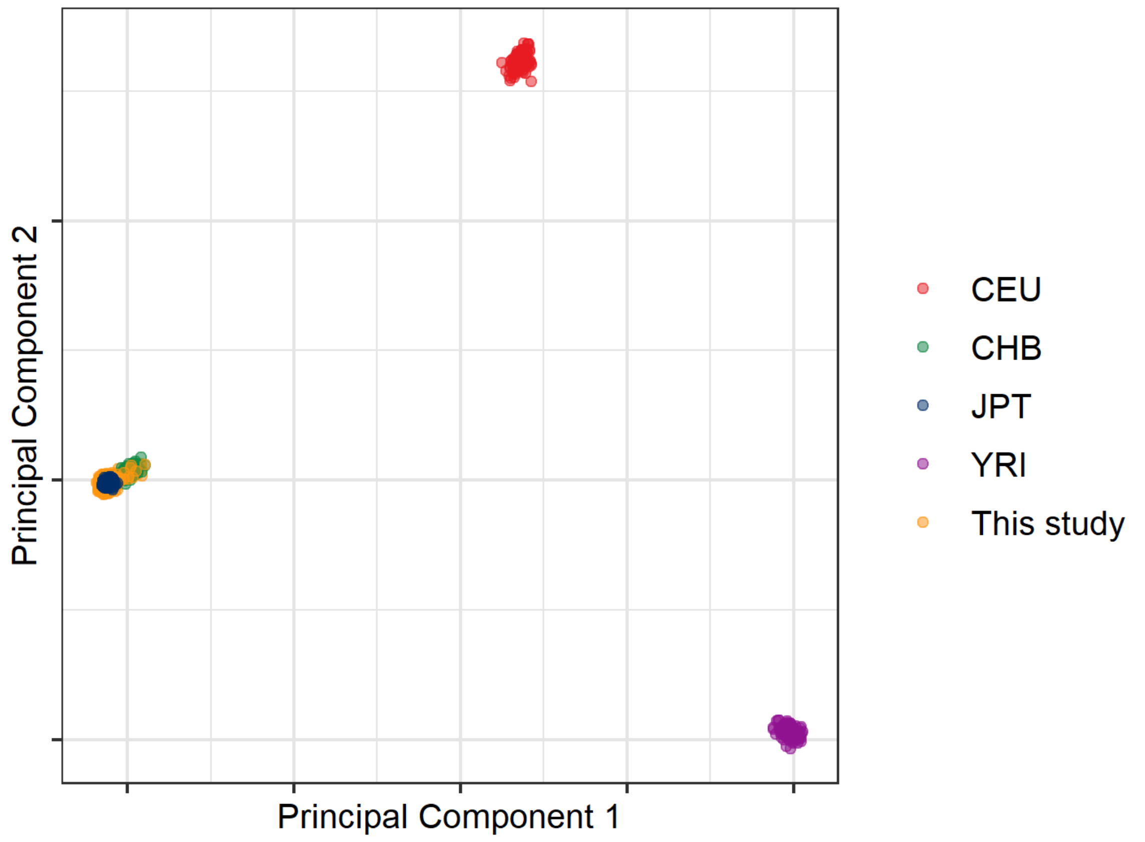


A principal component analysis (PCA) plot of the GWAS participants (COVID-19 cases and controls) along with International HapMap populations.

**Supplementary Figure 2. Manhattan and quantile-quantile plots of the GWAS**

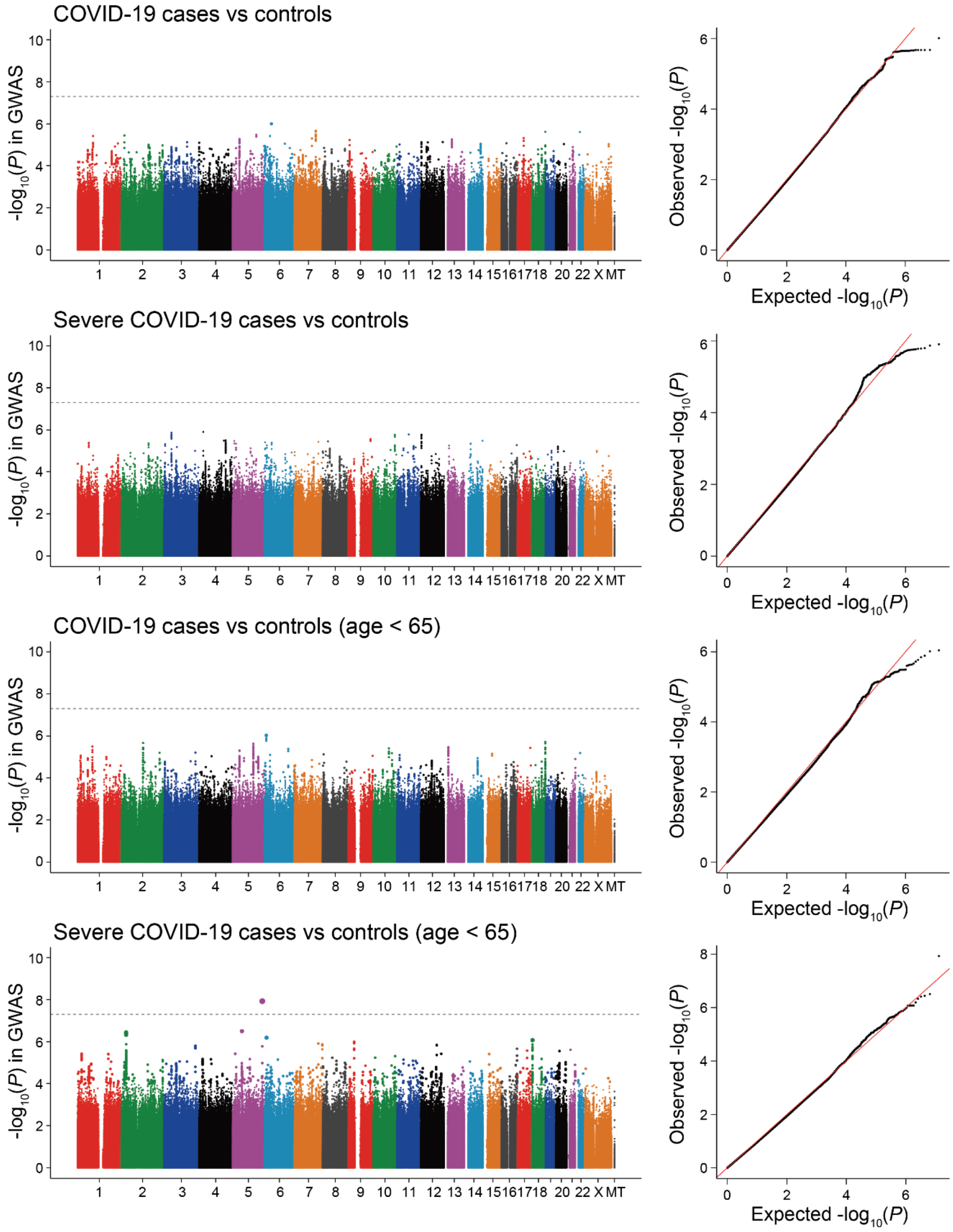


Manhattan plots and quantile-quantile plots of the Japanese GWAS of COVID-19. Dotted lines represent the genome-wide significance threshold of $P < 5.0 \times 10^{-8}$.

### Supplementary Figure 3. Regional association plots of the HLA imputation analysis

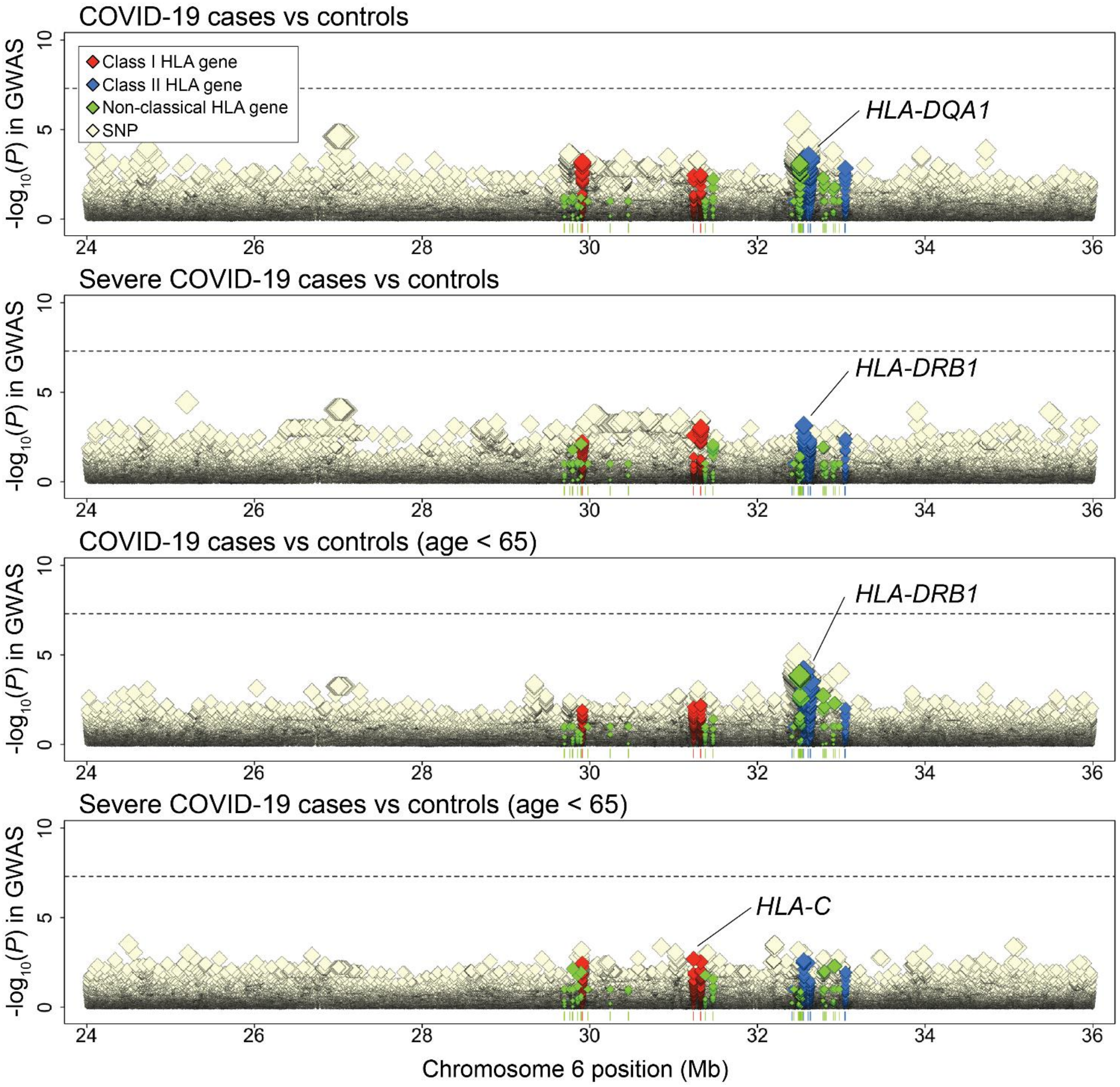


Regional association plots of the HLA imputation analysis results. Dots represent SNPs and HLA variants with colors according to the legend. Dotted lines represent the genome-wide significance threshold of $P < 5.0 \times 10^{-8}$. HLA genes with the most significant associations in each of the case-control phenotypes are indicated.

**Supplementary Table 1. A list of the medical institutes participating to Japan COVID-19 Task Force**

| |
|---|
| Aichi Cancer Center Hospital |
| Chiba University Graduate School of Medicine |
| Daini Osaka Police Hospital |
| Eiju General Hospital |
| Faculty of Medicine |
| Fujioka General Hospital |
| Fujisawa City Hospital |
| Fukujuji hospital |
| Fukuoka Tokushukai Hospital |
| Fukuoka University |
| Fukuoka University Hospital |
| Fukushima Medical University |
| Gifu University School of Medicine Graduate School of Medicine |
| Graduate School of Tokyo Institute of Technology |
| Gunma University Graduate School of Medicine |
| Himeji St. Mary's Hospital |
| International University of Health and Welfare Shioya Hospital |
| Ishikawa Prefectural Central Hospital |
| JA Toride medical hospital |
| Japan Community Health care Organization Kanazawa Hospital |
| Japanese Red Cross Medical Center |
| JCHO (Japan Community Health care Organization) Saitama Medical Center |
| Juntendo University Graduate School of Medicine |
| Kanagawa Cardiovascular and Respiratory Center |
| Kansai Electric Power Hospital |
| Kansai Medical University General Medical Center |
| Kansai Rosai Hospital |
| Kanto Rosai Hospital |
| Kawasaki Municipal Ida Hospital |
| Keio University Hospital |
| Keio University School of Medicine |
| Keiyu Hospital |
| KINSHUKAI Hanwa The Second Hospital |
| Kiryu Kosei General Hospital |
| Kitasato University |
| Kitasato University Kitasato Institute Hospital |
| KKR Sapporo Medical Center |
| Kumamoto City Hospital |
| Kurume University School of Medicine |
| Kyoto Prefectural University of Medicine |
| Kyoto University Graduate School of Medicine |

| |
|---|
| Kyushu University Graduate School of Medical Sciences |
| Matsumoto City Hostpital |
| Musashino Red Cross Hospital |
| Nagoya University Graduate School of Medicine |
| National Center for Global Health and Medicine |
| National Defense Medical College |
| National Hospital Organization Hokkaido Medical Center |
| National Hospital Organization Kumamoto Medical Center |
| National Hospital Organization Kyoto Medical Center |
| National Hospital Organization Kyushu Medical Center |
| National hospital organization Saitama Hospital |
| National Hospital Organization Tokyo Hospital |
| National Hospital Organization Tokyo Medical Center |
| NHO Kanazawa Medical Center |
| Nihon University School of Medicine |
| Niigata University |
| Okayama Rosai Hospital |
| Ome Municipal General Hospital |
| Osaka Saiseikai Nakatsu Hospital |
| Osaka University Graduate School of Medicine |
| Osaka University Hospital |
| Saiseikai Kumamoto Hospital |
| Saiseikai Utsunomiya Hospital |
| Saiseikai Yokohamashi Nanbu Hospital |
| Saitama Cardiovascular and Respiratory Center |
| Saitama City Hospital |
| Sano Kosei General Hospital |
| Sapporo City General Hospital |
| Showa University |
| Showa University Koto Toyosu Hospital |
| St Marianna University School of Medicine, Yokohama-City Seibu Hospital |
| St. Marianna University School of Medicine |
| St.Marianna University School of Medicine |
| Tachikawa Hospital |
| The Institute of Medical Science, the University of Tokyo |
| Tohoku University Graduate School of Medicine |
| Tokai University School of Medicine |
| Tokyo Medical and Dental University |
| Tokyo Medical and Dental University Hospital of Medicine |
| Tokyo Medical University Hospital |
| Tokyo Medical University Ibaraki Medical Center |
| Tokyo Metropolitan Police Hospital |
| Tokyo Saiseikai Central Hospital |
| Tokyo Women's Medical University Medical Center East |

| |
|---|
| Tokyo Medical and Dental University Hospital of Medicine |
| Tosei General Hospital |
| Toyohashi Municipal Hospital |
| Uji-Tokushukai Medical Center |
| University of Tsukuba |
| Yamagata University Faculty of Medicine |

Medical institutes and hospitals contributing to the current GWAS study are listed.

### Supplementary Table 2. Characteristics of the study participants

| Subjects | Age | Severity | No. subjects | Age (mean±SD) | Male proportion (%) |
|---|---|---|---|---|---|
| COVID-19 cases | All age | All | 2,393 | 56.0 ± 18.9 | 64.2 |
| | | Severe | 990 | 65.3 ± 13.9 | 73.9 |
| | | Non-severe | 1,391 | 49.3 ± 19.2 | 57.2 |
| | Age < 65 | All | 1,484 | 44.1 ± 13.3 | 65.7 |
| | | Severe | 440 | 52.6 ± 9.2 | 81.8 |
| | | Non-severe | 1,041 | 40.5 ± 13.1 | 58.9 |
| | Age ≥ 65 | All | 909 | 75.4 ± 7.1 | 61.7 |
| | | Severe | 550 | 75.5 ± 6.9 | 67.6 |
| | | Non-severe | 350 | 75.2 ± 7.6 | 52.3 |
| Controls | All Age | - | 3,289 | 53.1 ± 16.8 | 47.5 |
| | Age < 65 | - | 2,377 | 45.1 ± 12.1 | 47.8 |
| | Age ≥ 65 | - | 912 | 74.0 ± 6.2 | 46.5 |

**Supplementary Table 3. Detailed associations of the *DOCK2* rs60200309-A allele with COVID-19 susceptibility**

| Recruitment period | Age | Phenotype | No. subjects | | Risk allele frequency (A) | | OR (95%CI) | *P* |
|---|---|---|---|---|---|---|---|---|
| | | | Cases | Controls | Cases | Controls | | |
| April 2020 - Jan 2021 | All age | COVID-19 vs control | 2,393 | 3,289 | 0.119 | 0.101 | 1.24 (1.09-1.41) | 0.0011 |
| | | Severe COVID-19 vs control | 990 | 3,289 | 0.129 | 0.101 | 1.39 (1.16-1.66) | $3.1\times10^{-4}$ |
| | | Non-Severe COVID-19 vs control | 1,391 | 3,289 | 0.113 | 0.101 | 1.16 (1.00-1.35) | 0.054 |
| | | Severe COVID-19 vs Non-Severe COVID-19 | 990 | 1,391 | 0.129 | 0.113 | 1.27 (1.03-1.57) | 0.028 |
| | Age < 65 | COVID-19 vs control | 1,484 | 2,377 | 0.123 | 0.099 | 1.32 (1.13-1.55) | $5.1\times10^{-4}$ |
| | | Severe COVID-19 vs control | 440 | 2,377 | 0.159 | 0.099 | 2.01 (1.58-2.55) | $1.2\times10^{-8}$ |
| | | Non-Severe COVID-19 vs control | 1,041 | 2,377 | 0.107 | 0.099 | 1.08 (0.90-1.30) | 0.39 |
| | | Severe COVID-19 vs Non-Severe COVID-19 | 440 | 1,041 | 0.159 | 0.107 | 1.90 (1.43-2.52) | $1.1\times10^{-5}$ |
| | Age ≥ 65 | COVID-19 vs control | 909 | 912 | 0.113 | 0.106 | 1.08 (0.86-1.35) | 0.50 |
| | | Severe COVID-19 vs control | 550 | 912 | 0.104 | 0.106 | 0.93 (0.71-1.22) | 0.61 |
| | | Non-Severe COVID-19 vs control | 350 | 912 | 0.129 | 0.106 | 1.30 (0.98-1.73) | 0.069 |
| | | Severe COVID-19 vs Non-Severe COVID-19 | 550 | 350 | 0.104 | 0.129 | 0.75 (0.54-1.03) | 0.074 |
| April 2020 - July 2021 | Age < 65 | Severe COVID-19 vs control* | 96 | 792* | 0.189 | 0.101 | 2.38 (1.53-3.72) | $1.3\times10^{-5}$ |
| August 2020 - October 2021 | | | 165 | 792* | 0.148 | 0.096 | 1.79 (1.18-2.72) | $6.1\times10^{-3}$ |
| October 2020 - Jan 2021 | | | 179 | 793* | 0.154 | 0.099 | 1.92 (1.29-2.85) | $1.4\times10^{-3}$ |
| Meta-analysis | | | 440 | 2,377 | 0.159 | 0.099 | 2.00 (1.57-2.55) | $2.0\times10^{-8}$ |

* The controls were randomly split into the three groups in the association analysis stratified by the recruitment period.

**Supplementary Table 4. A replication study of the *DOCK2* variant in the pan-ancestry meta-analysis**

| Study | Ancestry | No. subjects | | Risk allele frequency (rs60200309-A) | OR (95%CI) | *P* |
|---|---|---|---|---|---|---|
| | | Cases | Controls | | | |
| UK Biobank | EUR | 1,824 | 428,622 | 0 | 0.44 (0.06-3.10) | 0.41 |
| | AFR | 83 | 9,193 | 0 | 0.22 (0.00-30.5) | 0.55 |
| | SAS | 71 | 10,252 | 0.00029 | 4.49 (0.51-39.6) | 0.18 |
| AncestryDNA | EUR | 636 | 72,097 | N.A. | 4.38 (1.10-17.5) | 0.037 |
| | AFR | 91 | 3,605 | N.A. | 0.60 (0.03-11.1) | 0.73 |
| | AMR | 104 | 7,653 | N.A. | 1.87 (0.83-4.23) | 0.13 |
| FinnGen | EUR | 82 | 238,353 | 0 | 0.35 (0.01-26.9) | 0.64 |
| Geisinger | EUR | 180 | 112,862 | 0 | 0.25 (0.00-30.0) | 0.57 |
| UPENN | AFR | 67 | 8,738 | 0 | 0.40 (0.00-43.3) | 0.70 |
| Meta-analysis | | 3,138 | 891,375 | 0.00082 | 1.73 (0.95-3.15) | 0.072 |

EUR; European, AFR; African, AMR; American.

**Supplementary Table 5. Meta-analysis of the discovery GWAS and the replication study of the *DOCK2* variant**

| rsID | Chr:position (b37) | Allele | Gene | Population | No. subjects | | Risk allele freq. (A) | | OR (95%CI) | *P* |
|---|---|---|---|---|---|---|---|---|---|---|
| | | | | | Cases | Controls | Cases | Controls | | |
| rs60200309 | 5:169519612 | G/A | *DOCK2* | Japanese | 440 | 2,377 | 0.159 | 0.099 | 2.01 (1.58-2.55) | $1.2\times10^{-8}$ |
| | | | | Pan-ancestry | 3,138 | 891,375 | 0.0026 | 0.0008 | 1.73 (0.95-3.15) | 0.072 |
| | | | | Meta-analysis | - | - | - | - | 1.97 (1.57-2.46) | $1.2\times10^{-9}$ |

### Supplementary Table 6. Associations of the previously reported COVID-19 risk variants

| rsID | Chr:position (b37) | Allele | Gene | Phenotype | Risk allele frequency | | OR (95%CI) | *P* |
|---|---|---|---|---|---|---|---|---|
| | | Risk/non-risk | | | Case | Control | | |
| rs2271616 | 3:45838013 | T/G | *SLC6A20* | COVID-19 vs control | 0.128 | 0.129 | 0.98 (0.87-1.10) | 0.75 |
| | | | | Severe COVID-19 vs control | 0.129 | 0.129 | 0.97 (0.81-1.14) | 0.68 |
| | | | | COVID-19 vs control (age<65) | 0.129 | 0.129 | 1.01 (0.87-1.16) | 0.92 |
| | | | | Severe COVID-19 vs control (age<65) | 0.134 | 0.129 | 0.97 (0.77-1.23) | 0.79 |
| rs35081325 | 3:45889921 | T/A | *LZTFL1* | COVID-19 vs control | 0.0021 | 0.0013 | 2.49 (0.77-8.08) | 0.13 |
| | | | | Severe COVID-19 vs control | 0.0032 | 0.0013 | **7.06 (1.63-30.6)** | **0.0090** |
| | | | | COVID-19 vs control (age<65) | 0.0021 | 0.0013 | 2.22 (0.53-9.39) | 0.28 |
| | | | | Severe COVID-19 vs control (age<65) | 0.0030 | 0.0013 | **11.8 (1.64-85.5)** | **0.014** |
| rs11919389 | 3:101424458 | T/C | *RPL24* | COVID-19 vs control | 0.626 | 0.625 | 1.02 (0.94-1.10) | 0.66 |
| | | | | Severe COVID-19 vs control | 0.619 | 0.625 | 1.02 (0.91-1.14) | 0.79 |
| | | | | COVID-19 vs control (age<65) | 0.623 | 0.632 | 0.97 (0.88-1.07) | 0.56 |
| | | | | Severe COVID-19 vs control (age<65) | 0.615 | 0.632 | 0.95 (0.81-1.12) | 0.57 |
| rs1886814 | 6:41502683 | C/A | *FOXP4* | COVID-19 vs control | 0.307 | 0.285 | **1.14 (1.04-1.24)** | **0.0035** |
| | | | | Severe COVID-19 vs control | 0.317 | 0.285 | **1.29 (1.13-1.46)** | **$9.1 \times 10^{-5}$** |
| | | | | COVID-19 vs control (age<65) | 0.314 | 0.285 | **1.17 (1.05-1.30)** | **0.0045** |
| | | | | Severe COVID-19 vs control (age<65) | 0.340 | 0.285 | **1.42 (1.19-1.69)** | **$1.2 \times 10^{-4}$** |
| rs72711165 | 8:125336564 | C/T | *TMEM65* | COVID-19 vs control | 0.035 | 0.031 | 1.15 (0.93-1.42) | 0.21 |
| | | | | Severe COVID-19 vs control | 0.034 | 0.031 | 1.15 (0.85-1.57) | 0.37 |
| | | | | COVID-19 vs control (age<65) | 0.040 | 0.030 | **1.31 (1.01-1.70)** | **0.040** |
| | | | | Severe COVID-19 vs control (age<65) | 0.044 | 0.030 | 1.42 (0.94-2.16) | 0.094 |
| rs529565 | 9:136149500 | C/T | *ABO* | COVID-19 vs control | 0.475 | 0.456 | 1.08 (1.00-1.16) | 0.057 |
| | | | | Severe COVID-19 vs control | 0.494 | 0.456 | **1.18 (1.06-1.32)** | **0.0028** |
| | | | | COVID-19 vs control (age<65) | 0.476 | 0.458 | 1.08 (0.99-1.19) | 0.089 |
| | | | | Severe COVID-19 vs control (age<65) | 0.510 | 0.458 | **1.22 (1.04-1.42)** | **0.012** |
| rs10774671 | 12:113357193 | A/G | *OAS1* | COVID-19 vs control | 0.800 | 0.791 | 1.06 (0.96-1.17) | 0.24 |
| | | | | Severe COVID-19 vs control | 0.802 | 0.791 | 1.08 (0.94-1.24) | 0.30 |
| | | | | COVID-19 vs control (age<65) | 0.805 | 0.794 | 1.07 (0.95-1.20) | 0.28 |
| | | | | Severe COVID-19 vs control (age<65) | 0.814 | 0.794 | 1.09 (0.89-1.34) | 0.38 |
| rs1819040 | 17:44219831 | T/A | *KANSL1* | Not imputable | | | | |
| rs77534576 | 17:47940666 | T/C | *TAC4* | COVID-19 vs control | 0.048 | 0.045 | 1.07 (0.89-1.29) | 0.46 |
| | | | | Severe COVID-19 vs control | 0.057 | 0.045 | **1.29 (1.01-1.65)** | **0.043** |
| | | | | COVID-19 vs control (age<65) | 0.051 | 0.044 | 1.16 (0.93-1.45) | 0.20 |
| | | | | Severe COVID-19 vs control (age<65) | 0.060 | 0.044 | 1.33 (0.95-1.88) | 0.098 |
| rs2109069 | 19:4719443 | A/G | *DPP9* | COVID-19 vs control | 0.123 | 0.111 | 1.10 (0.98-1.24) | 0.12 |
| | | | | Severe COVID-19 vs control | 0.133 | 0.111 | **1.20 (1.02-1.42)** | **0.031** |
| | | | | COVID-19 vs control (age<65) | 0.120 | 0.113 | 1.06 (0.91-1.22) | 0.46 |
| | | | | Severe COVID-19 vs control (age<65) | 0.131 | 0.113 | 1.11 (0.88-1.41) | 0.39 |
| rs74956615 | 19:10427721 | A/T | *RAVER1* | Not imputable | | | | |
| rs4801778 | 19:49370609 | G/T | *PLEKHA4* | COVID-19 vs control | 0.978 | 0.975 | 1.15 (0.88-1.49) | 0.30 |
| | | | | Severe COVID-19 vs control | 0.977 | 0.975 | 1.06 (0.73-1.54) | 0.77 |
| | | | | COVID-19 vs control (age<65) | 0.978 | 0.973 | 1.23 (0.89-1.69) | 0.20 |
| | | | | Severe COVID-19 vs control (age<65) | 0.980 | 0.973 | 1.29 (0.74-2.24) | 0.37 |
| rs13050728 | 21:34615210 | T/C | *IFNAR2* | COVID-19 vs control | 0.573 | 0.549 | **1.10 (1.02-1.19)** | **0.015** |
| | | | | Severe COVID-19 vs control | 0.572 | 0.549 | 1.08 (0.97-1.22) | 0.17 |
| | | | | COVID-19 vs control (age<65) | 0.580 | 0.543 | **1.16 (1.06-1.28)** | **0.0024** |
| | | | | Severe COVID-19 vs control (age<65) | 0.605 | 0.543 | **1.28 (1.08-1.50)** | **0.0039** |

Associations with *P* < 0.05 are highlighted in bold.

**Supplementary Table 7. HLA variant associations with COVID-19 risk**

(Results are indicated in a separate Microsoft Excel file)

### Supplementary Table 8. Associations of the ABO blood type with COVID-19 risk

#### Associations of the target blood type compared with other blood types

| Age | Phenotype | Target vs other blood types | Target blood type freq. | | OR (95%CI) | *P* |
|---|---|---|---|---|---|---|
| | | | Cases | control | | |
| All age | COVID-19 vs control | A vs AB/B/O | 0.411 | 0.390 | 1.10 (0.98-1.23) | 0.093 |
| | | B vs A/AB/O | 0.213 | 0.218 | 0.95 (0.83-1.09) | 0.46 |
| | | AB vs A/B/O | 0.106 | 0.095 | 1.13 (0.95-1.35) | 0.18 |
| | | O vs A/AB/B | 0.270 | 0.297 | **0.88 (0.78-1.00)** | **0.041** |
| | Severe COVID-19 vs control | A vs AB/B/O | 0.407 | 0.390 | 1.04 (0.89-1.22) | 0.61 |
| | | B vs A/AB/O | 0.221 | 0.218 | 1.00 (0.83-1.21) | 0.96 |
| | | AB vs A/B/O | 0.120 | 0.095 | **1.41 (1.10-1.81)** | **0.0065** |
| | | O vs A/AB/B | 0.253 | 0.297 | **0.81 (0.68-0.96)** | **0.016** |
| Age < 65 | COVID-19 vs control | A vs AB/B/O | 0.412 | 0.381 | **1.18 (1.03-1.35)** | **0.017** |
| | | B vs A/AB/O | 0.214 | 0.222 | 0.92 (0.78-1.08) | 0.33 |
| | | AB vs A/B/O | 0.109 | 0.099 | 1.09 (0.88-1.36) | 0.41 |
| | | O vs A/AB/B | 0.265 | 0.298 | **0.85 (0.73-0.98)** | **0.027** |
| | Severe COVID-19 vs control | A vs AB/B/O | 0.394 | 0.381 | 1.08 (0.86-1.35) | 0.52 |
| | | B vs A/AB/O | 0.235 | 0.222 | 1.06 (0.81-1.37) | 0.67 |
| | | AB vs A/B/O | 0.137 | 0.099 | **1.40 (1.00-1.94)** | **0.048** |
| | | O vs A/AB/B | 0.235 | 0.298 | **0.73 (0.56-0.93)** | **0.014** |

#### Associations of the target blood type compared with the O blood type

| Age | Phenotype | Target blood types vs O | Target blood type freq. | | OR (95%CI) | *P* |
|---|---|---|---|---|---|---|
| | | | Cases | control | | |
| All age | COVID-19 vs control | A vs O | 0.411 | 0.390 | **1.16 (1.02-1.32)** | **0.029** |
| | | B vs O | 0.213 | 0.218 | 1.05 (0.90-1.23) | 0.52 |
| | | AB vs O | 0.106 | 0.095 | **1.22 (1.00-1.48)** | **0.050** |
| | Severe COVID-19 vs control | A vs O | 0.407 | 0.390 | 1.20 (0.99-1.46) | 0.068 |
| | | B vs O | 0.221 | 0.218 | 1.17 (0.93-1.46) | 0.18 |
| | | AB vs O | 0.120 | 0.095 | **1.57 (1.19-2.08)** | **0.0015** |
| Age < 65 | COVID-19 vs control | A vs O | 0.412 | 0.381 | **1.25 (1.06-1.48)** | **0.0074** |
| | | B vs O | 0.214 | 0.222 | 1.06 (0.88-1.28) | 0.55 |
| | | AB vs O | 0.109 | 0.099 | 1.23 (0.96-1.56) | 0.097 |
| | Severe COVID-19 vs control | A vs O | 0.394 | 0.381 | 1.32 (0.99-1.76) | 0.057 |
| | | B vs O | 0.235 | 0.222 | 1.32 (0.96-1.82) | 0.084 |
| | | AB vs O | 0.137 | 0.099 | **1.67 (1.14-2.44)** | **0.0081** |

#### Associations of the target blood type compared with the A blood type

| Age | Phenotype | Target blood types vs A | Target blood type freq. | | OR (95%CI) | *P* |
|---|---|---|---|---|---|---|
| | | | Cases | control | | |
| All age | COVID-19 vs control | B vs A | 0.213 | 0.218 | 0.91 (0.79-1.05) | 0.21 |
| | | AB vs A | 0.106 | 0.095 | 1.06 (0.87-1.28) | 0.57 |
| | | O vs A | 0.270 | 0.297 | **0.86 (0.76-0.98)** | **0.029** |

| | | | | | | |
|---|---|---|---|---|---|---|
| | Severe COVID-19 vs control | B vs A | 0.221 | 0.218 | 0.98 (0.80-1.20) | 0.85 |
| | | AB vs A | 0.120 | 0.095 | **1.34 (1.03-1.75)** | **0.029** |
| | | O vs A | 0.253 | 0.297 | 0.83 (0.69-1.01) | 0.068 |
| Age < 65 | COVID-19 vs control | B vs A | 0.214 | 0.222 | 0.85 (0.71-1.02) | 0.079 |
| | | AB vs A | 0.109 | 0.099 | 0.98 (0.78-1.23) | 0.87 |
| | | O vs A | 0.265 | 0.298 | **0.80 (0.68-0.94)** | **0.0074** |
| | Severe COVID-19 vs control | B vs A | 0.235 | 0.222 | 1.01 (0.75-1.35) | 0.96 |
| | | AB vs A | 0.137 | 0.099 | 1.34 (0.93-1.91) | 0.12 |
| | | O vs A | 0.235 | 0.298 | 0.76 (0.57-1.01) | 0.057 |

Associations of the target blood type compared with the B blood type

| Age | Phenotype | Target blood types vs B | Target blood type freq. | | OR (95%CI) | *P* |
|---|---|---|---|---|---|---|
| | | | Cases | control | | |
| All age | COVID-19 vs control | A vs B | 0.411 | 0.390 | 1.10 (0.95-1.27) | 0.21 |
| | | AB vs B | 0.106 | 0.095 | 1.16 (0.94-1.43) | 0.16 |
| | | O vs B | 0.270 | 0.297 | 0.95 (0.81-1.11) | 0.52 |
| | Severe COVID-19 vs control | A vs B | 0.407 | 0.390 | 1.02 (0.83-1.26) | 0.85 |
| | | AB vs B | 0.120 | 0.095 | **1.35 (1.01-1.80)** | **0.041** |
| | | O vs B | 0.253 | 0.297 | 0.86 (0.69-1.07) | 0.18 |
| Age < 65 | COVID-19 vs control | A vs B | 0.412 | 0.381 | 1.17 (0.98-1.40) | 0.079 |
| | | AB vs B | 0.109 | 0.099 | 1.15 (0.89-1.47) | 0.29 |
| | | O vs B | 0.265 | 0.298 | 0.94 (0.78-1.14) | 0.55 |
| | Severe COVID-19 vs control | A vs B | 0.394 | 0.381 | 0.99 (0.74-1.33) | 0.96 |
| | | AB vs B | 0.137 | 0.099 | 1.25 (0.85-1.84) | 0.26 |
| | | O vs B | 0.235 | 0.298 | 0.76 (0.55-1.04) | 0.084 |

Associations of the target blood type compared with the A or B blood types

| Age | Phenotype | Target blood types vs A or B | Target blood type freq. | | OR (95%CI) | *P* |
|---|---|---|---|---|---|---|
| | | | Cases | control | | |
| All age | COVID-19 vs control | AB vs A or B | 0.106 | 0.095 | 1.09 (0.91-1.31) | 0.34 |
| | | O vs A or B | 0.270 | 0.297 | 0.89 (0.79-1.01) | 0.072 |
| | Severe COVID-19 vs control | AB vs A or B | 0.120 | 0.095 | **1.34 (1.04-1.73)** | **0.023** |
| | | O vs A or B | 0.253 | 0.297 | 0.84 (0.70-1.01) | 0.058 |
| Age < 65 | COVID-19 vs control | AB vs A or B | 0.109 | 0.099 | 1.04 (0.83-1.30) | 0.73 |
| | | O vs A or B | 0.265 | 0.298 | **0.85 (0.73-0.99)** | **0.035** |
| | Severe COVID-19 vs control | AB vs A or B | 0.137 | 0.099 | 1.29 (0.91-1.81) | 0.14 |
| | | O vs A or B | 0.235 | 0.298 | **0.75 (0.58-0.98)** | **0.034** |

Associations with *P* < 0.05 are highlighted in bold.

**Supplementary Table 9. Details of the GWAS studies of the exposure phenotypes in the Mendelian randomization analysis**

| Population | Phenotype | No. subjects | | | Reference | PMID |
|---|---|---|---|---|---|---|
| | | Cases | Controls | Total | | |
| Japanese | Body mass index (BMI) | - | - | 173,430 | Akiyama et al. *Nat Genet* (2017) | 28892062 |
| | Type 2 diabetes (T2D) | 77,418 | 356,122 | 433,540 | Spracklen et al. *Nature* (2020) | 32499647 |
| | Cigarettes per day (CPD) | - | - | 72,655 | Matoba et al. Nat Hum Behav(2019) | 31089300 |
| | Asthma | 8,216 | 201,592 | 209,808 | Ishigaki et al. *Nat Genet* (2020) | 32514122 |
| | Systolic blood pressure (sBP) | - | - | 183,785 | Takeuchi et al. *Nat Commun* (2018) | 30487518 |
| | Diastolic blood pressure (dBP) | - | - | 183,785 | Takeuchi et al. *Nat Commun* (2018) | 30487518 |
| | Estimated glomerular filtration rate (eGFR) | - | - | 143,658 | Kanai et al. *Nat Genet* (2018) | 29403010 |
| | Serum uric acids (UA) | - | - | 121,745 | Nakatochi et al. *Commun Biol* (2019) | 30993211 |
| | Gout | 3,053 | 4,554 | 7,607 | Nakayama et al. *Ann Rheum Dis* (2020) | 32238385 |
| | Rheumatoid arthritis (RA) | 4,873 | 17,641 | 22,514 | Okada et al. *Nature* (2014) | 24390342 |
| | Systemic lupus erythematosus (SLE) | 13,377 | 194,993 | 208,370 | Yin et al. *Ann Rheum Dis* (2020) | 33272962 |
| European | Body mass index (BMI) | - | - | 681,275 | Yengo et al. *Hum Mol Genet* (2018) | 30124842 |
| | Type 2 diabetes (T2D) | 148,726 | 965,732 | 1,114,458 | Vujkovic et al. *Nat Gene*t (2020) | 32541925 |
| | Cigarettes per day (CPD) | - | - | 337,334 | Liu et al. Nat Genet (2019) | 30643251 |
| | Asthma | 64,538 | 329,321 | 393,859 | Han et al. *Nat Commun* (2020) | 32296059 |
| | Systolic blood pressure (sBP) | - | - | 757,601 | Evangelou et al. *Nat Genet* (2018) | 30224653 |
| | Diastolic blood pressure (dBP) | - | - | 757,601 | Evangelou et al. *Nat Genet* (2018) | 30224653 |
| | Estimated glomerular filtration rate (eGFR) | - | - | 567,460 | Wuttke et al. *Nat Genet* (2019) | 31152163 |
| | Serum uric acids (UA) | - | - | 288,649 | Tin et al. *Nat Genet* (2019) | 31578528 |
| | Gout | 13,179 | 750,634 | 763,813 | Tin et al. *Nat Genet* (2019) | 31578528 |
| | Rheumatoid arthritis (RA) | 14,361 | 43,923 | 58,284 | Okada et al. *Nature* (2014) | 24390342 |
| | Systemic lupus erythematosus (SLE) | 5,201 | 9,066 | 14,267 | Bentham et al. *Nat Genet* (2015) | 26502338 |

### Supplementary Table 10. Results of the cross-population Mendelian randomization analysis on COVID-19

| Exposure phenotypes (Japanese) | COVID-19 vs control | | | | Severe COVID-19 vs control | | | | COVID-19 vs control (age<65) | | | | Severe COVID-19 vs control (age<65) | | | |
|---|---|---|---|---|---|---|---|---|---|---|---|---|---|---|---|---|
| | $N_{SNP}$ | Beta | SE | *P* | $N_{SNP}$ | Beta | SE | *P* | $N_{SNP}$ | Beta | SE | *P* | $N_{SNP}$ | Beta | SE | *P* |
| Body mass index (BMI) | 76 | 0.296 | 0.173 | 0.087 | 76 | **0.676** | **0.249** | **0.0067** | 76 | 0.380 | 0.213 | 0.075 | 76 | **0.946** | **0.353** | **0.0074** |
| Type 2 diabetes (T2D) | 164 | 0.050 | 0.048 | 0.30 | 164 | 0.075 | 0.069 | 0.27 | 164 | 0.032 | 0.059 | 0.59 | 164 | 0.069 | 0.099 | 0.48 |
| Cigarettes per day (CPD) | 5 | 0.167 | 0.310 | 0.59 | 5 | -0.236 | 0.449 | 0.60 | 5 | 0.015 | 0.381 | 0.97 | 5 | -0.887 | 0.637 | 0.16 |
| Asthma | 7 | **0.376** | **0.137** | **0.0061** | 7 | 0.165 | 0.197 | 0.40 | 7 | **0.401** | **0.169** | **0.018** | 7 | 0.195 | 0.279 | 0.49 |
| Systolic blood pressure (sBP) | 54 | 0.011 | 0.011 | 0.35 | 54 | -0.008 | 0.016 | 0.62 | 54 | 0.014 | 0.014 | 0.31 | 54 | -0.022 | 0.023 | 0.33 |
| Diastolic blood pressure (dBP) | 41 | 0.017 | 0.021 | 0.42 | 41 | -0.022 | 0.030 | 0.46 | 41 | 0.014 | 0.025 | 0.57 | 41 | -0.051 | 0.042 | 0.23 |
| Estimated glomerular filtration rate (eGFR) | 67 | -0.070 | 0.170 | 0.68 | 67 | 0.082 | 0.245 | 0.74 | 67 | 0.027 | 0.209 | 0.90 | 67 | 0.147 | 0.347 | 0.67 |
| Serum uric acids (UA) | 36 | 0.163 | 0.095 | 0.088 | 36 | 0.024 | 0.137 | 0.86 | 36 | **0.279** | **0.119** | **0.019** | 36 | 0.216 | 0.199 | 0.28 |
| Gout | 10 | **0.078** | **0.028** | **0.0048** | 10 | 0.018 | 0.039 | 0.65 | 10 | **0.104** | **0.035** | **0.0027** | 10 | 0.059 | 0.057 | 0.30 |
| Rheumatoid arthritis (RA) | 15 | -0.065 | 0.048 | 0.18 | 15 | -0.018 | 0.070 | 0.80 | 15 | -0.102 | 0.059 | 0.083 | 15 | -0.067 | 0.097 | 0.49 |
| Systemic lupus erythematosus (SLE) | 100 | **-0.087** | **0.027** | **0.0014** | 100 | -0.060 | 0.039 | 0.12 | 100 | -0.058 | 0.033 | 0.084 | 100 | 0.022 | 0.056 | 0.70 |

| Exposure phenotypes (European) | Self-reported COVID-19 (C2) | | | | Hospitalized COVID-19 (B2) | | | | Severe COVID-19 (A2) | | | |
|---|---|---|---|---|---|---|---|---|---|---|---|---|
| | $N_{SNP}$ | Beta | SE | *P* | $N_{SNP}$ | Beta | SE | *P* | $N_{SNP}$ | Beta | SE | *P* |
| Body mass index (BMI) | 486 | **0.175** | **0.030** | **$8.5\times10^{-9}$** | 486 | **0.398** | **0.060** | **$3.2\times10^{-11}$** | 486 | **0.405** | **0.090** | **$6.2\times10^{-6}$** |
| Type 2 diabetes (T2D) | 414 | **0.027** | **0.012** | **0.019** | 413 | **0.061** | **0.023** | **0.0078** | 417 | 0.068 | 0.035 | 0.050 |
| Cigarettes per day (CPD) | 39 | 0.032 | 0.067 | 0.63 | 39 | 0.255 | 0.132 | 0.054 | 39 | 0.096 | 0.199 | 0.63 |
| Asthma | 124 | -0.026 | 0.017 | 0.14 | 119 | -0.025 | 0.035 | 0.48 | 124 | -0.059 | 0.052 | 0.26 |
| Systolic blood pressure (sBP) | 396 | -0.0015 | 0.0018 | 0.41 | 394 | 0.0019 | 0.0036 | 0.59 | 396 | -0.0036 | 0.0055 | 0.51 |
| Diastolic blood pressure (dBP) | 408 | -0.0038 | 0.0030 | 0.20 | 405 | -0.0021 | 0.0061 | 0.72 | 407 | -0.011 | 0.009 | 0.25 |
| Estimated glomerular filtration rate (eGFR) | 256 | 0.014 | 0.211 | 0.95 | 255 | -0.459 | 0.423 | 0.28 | 256 | **-1.315** | **0.649** | **0.043** |
| Serum uric acids (UA) | 121 | -0.013 | 0.019 | 0.51 | 121 | 0.028 | 0.038 | 0.46 | 122 | 0.048 | 0.056 | 0.39 |
| Gout | 27 | -0.007 | 0.012 | 0.58 | 27 | -0.020 | 0.024 | 0.42 | 28 | -0.009 | 0.036 | 0.81 |
| Rheumatoid arthritis (RA) | 63 | -0.010 | 0.011 | 0.37 | 63 | -0.028 | 0.022 | 0.20 | 63 | 0.028 | 0.033 | 0.40 |
| Systemic lupus erythematosus (SLE) | 41 | -0.011 | 0.007 | 0.11 | 41 | -0.016 | 0.014 | 0.25 | 41 | -0.024 | 0.020 | 0.24 |
